# The Potential Clinical and Economic Impact of a Combination COVID-19 and Influenza Vaccine (mRNA-1083) in Canada

**DOI:** 10.64898/2026.05.18.26353482

**Authors:** Kelly Fust, Michele Kohli, Shannon Cartier, Nicolas Van de Velde, Darshan Mehta, Michelle Blake

## Abstract

**Aims:** COVID-19 and influenza continue to impose a substantial burden on the Canadian healthcare system, particularly among adults aged ≥65 years. This study compared the clinical and economic outcomes of a “Stand-alone” vaccination strategy with separate influenza and COVID-19 vaccines versus a “Combination” strategy incorporating mRNA-1083, an investigational vaccine targeting both infections.

**Methods:** The study adopted the public healthcare payer perspective and adapted a previously published static model to predict COVID-19 and influenza infections across a one-year time horizon. Relative vaccine effectiveness (rVE) for mRNA-1083 against COVID-19 compared with the stand-alone vaccine (SPIKEVAX ®) was based on the pivotal clinical trial of mRNA-1083’s COVID-19 component (mRNA-1283). For influenza, no incremental VE was assumed versus the adjuvanted stand-alone vaccine (FLUAD®). Infections were modeled independently. Clinical outcomes included symptomatic infections, hospitalizations, and deaths. The economically justifiable price (EJP) was calculated at the willingness-to-pay (WTP) threshold of $50,000 per quality-adjusted life-year (QALY) gained. mRNA-1083 uptake was assumed to yield absolute increases in COVID-19 and influenza coverage by 10% and 3%, respectively.

**Results:** Compared with the Stand-alone strategy, the Combination strategy was projected to reduce the number of COVID-19-related symptomatic infections, hospitalizations, and deaths (n=71,074; 5,008; 935, respectively), and corresponding influenza outcomes (n=3,985; 362; 69, respectively). The use of mRNA-1083 within the Combination strategy generated a cost-savings of $90,440,471 in vaccine administration fees and an EJP of $304 per dose. Results were sensitive to rVE, coverage, administration fees, mortality and incidence.

**Limitations:** mRNA-1083’s rVE is being evaluated in clinical trials and the impact of mRNA-1083 on vaccine coverage and administration fees is uncertain.

**Conclusions:** mRNA-1083 may reduce the burden of COVID-19 and influenza in adults aged ≥65 years in Canada, while offering good economic value because it has the potential to increase coverage and VE while reducing administration fees.

## Introduction

Despite established seasonal vaccination programs targeting influenza and severe acute respiratory syndrome coronavirus 2 (SARS-CoV-2; [COVID-19]) in Canada, these respiratory infections continue to place a substantial burden on the healthcare system, disproportionately affecting adults aged ≥65 years [1]. Surveillance data from FluWatch+, Canada’s national respiratory virus surveillance system coordinated by the Public Health Agency of Canada (PHAC), indicate that from August 24, 2024 to August 23, 2025, there were 25,625 influenza-associated hospitalizations (76.9 per 100,000 population; 315.2 per 100,000 among individuals aged ≥65 years), including 728 intensive care unit (ICU) admissions and 697 deaths associated with influenza [2,3]. During the same period, 33,755 COVID-19 associated hospitalizations were reported (101.3 per 100,000 population; 506.5 per 100,000 among those ≥65 years), including 469 ICU admissions and 2,315 COVID-19-attributable deaths, 93% of which occurred among individuals aged ≥65 years [1]. COVID-19 has also been associated with longer-term sequelae, such as post-COVID condition and exacerbation of underlying medical conditions (e.g., increased risk of cardiovascular outcomes in those with chronic respiratory disease) [4–9].

Although SARS-CoV-2 activity fluctuates year-round, cases occurring in the fall and winter that coincide with seasonal influenza circulation place additional pressure on healthcare services [10,11]. Together, these viruses contribute to significant morbidity and healthcare resource utilization in Canada, particularly among older adults, underscoring the continued importance of evolving toward effective and efficient seasonal vaccination strategies [12,13].

In Canada, the populations recommended for influenza and COVID-19 vaccination by the National Advisory Committee on Immunization (NACI) largely overlap [10,11]. In its 2025-2026 and 2026-2027 seasonal influenza vaccine statements, NACI recommends influenza vaccination in the fall for all individuals aged ≥6 months, with particular emphasis on groups at higher risk of severe disease, including adults aged ≥65 years, as soon as the vaccine is available [14,15]. Similarly, NACI recommends an updated COVID-19 vaccine during the fall vaccination campaign for the 2025-2026 season for adults aged ≥65 years [16].

Given the overlap in target populations and seasonal disease burden, co-administration of influenza and COVID-19 vaccines is routinely practiced and recommended across provinces [17–19]. For example, Ontario Ministry of Health guidance for the 2025/2026 respiratory season encourages early influenza immunization early in the fall among high-risk populations (including those aged ≥65 years) to optimize concomitant immunization with the updated COVID-19 vaccine [17]. Based on evidence indicating no meaningful concerns regarding the efficacy, safety, or immunogenicity of concurrent administration, NACI has identified co-administration of these vaccines as an approach to improve vaccine delivery efficiency and vaccine uptake [10].

Despite these recommendations, vaccine coverage rates (VCR) for influenza and COVID-19 remain suboptimal among older adults in Canada [20–22]. During the 2024-2025 season, 63% of adults aged ≥65 years received the influenza vaccine, compared with 54% for COVID-19 [23]. Recent surveys of older Canadians suggest that logistical and informational barriers, such as difficulty booking appointments and the lack of concomitant vaccine availability, limit access to influenza and COVID-19 vaccines [21,24,25]. Also, differences in who administers these vaccines may hinder the feasibility of co-administration. For example, during the 2025-2026 season in Ontario, all publicly listed pharmacy vaccination sites (n=3,923) offered influenza vaccination, whereas only approximately half (n=1,972) offered COVID-19 vaccination, indicating that access to both vaccines within a single setting was not consistently available [26].

The gap between current coverage and recommendations highlights the need for more efficient vaccination strategies, for which combination vaccines targeting multiple respiratory viruses may offer a solution [27]. By enabling protection against both influenza and COVID-19 within a single administration, they may simplify vaccine delivery, reduce reliance on coadministration, and improve the efficiency of seasonal vaccination programs [28]. These advantages may help optimize uptake and timing of vaccination in priority populations, while also generating cost-and time-savings within the healthcare system [29–33]. The population health impact, economic value and appropriate pricing of an influenza-COVID-19 combination vaccine are important considerations for program planning and reimbursement decisions, but have not been characterized for Canada.

mRNA-1083 is an investigational mRNA vaccine designed to provide protection against influenza and COVID-19 in a single administration and, pending regulatory approval, may represent the first combination vaccine targeting respiratory viral infections available in Canada [34]. mRNA-1083 is anticipated to be indicated for the active immunization for the prevention of influenza and COVID-19 in adults aged ≥50 years [35]. mRNA-1083 combines components from the influenza vaccine candidate mRNA-1010 (currently under Health Canada review) and the next-generation COVID-19 vaccine, mRNA-1283 (mNEXSPIKE™, approved by Health Canada in 2025) [36]. mRNA-1083 was approved in April 2026 by the European Medicines Agency [37]. In a pivotal phase 3 randomized clinical trial in adults aged ≥50 years, mRNA-1083 demonstrated non-inferior immunogenicity compared with co-administered licensed influenza and COVID-19 vaccines across all vaccine-matched strains [34]. Among adults aged ≥65 years, mRNA-1083 met pre-specified non-inferiority criteria versus co-administered high-dose quadrivalent and COVID-19 vaccinations across all influenza and SARS-CoV-2 strains (GMR lower bound >0.667; seroconversion / sero-response difference >−10%), with higher immune responses observed for three clinically relevant influenza strains (A/H1N1, A/H3N2, B/Victoria) and for SARS-CoV-2, and an acceptable safety profile [34].

NACI preferentially recommends enhanced influenza vaccines that provide stronger protection for use by those ≥65 years, with Fluad® (adjuvanted standard dose influenza vaccine; CSL Seqirus [IIV-Adj]) and Fluzone-HD® (high dose influenza vaccine; Sanofi Pasteur [IIV-HD]) being the most commonly used in Canada [14,15]. These “egg-based” vaccines are manufactured by replicating the virus within chicken eggs. Viruses can adapt to this process because they develop mutations that allow faster replication [38]. If this “egg-adaptation” affects the antigenic match to circulating viruses, it may also lead to a reduction in the effectiveness of the vaccine [39]. Data from observational studies comparing egg-based vaccines to ones replicated using alternative measures have demonstrated superior VE across multiple seasons [40,41]. While egg-adaptation may affect both IIV-Adj and IIV-HD, it will not impact mRNA-1083.

This study aimed to estimate the clinical and economic impact of two vaccination allocation strategies in adults aged ≥65 years: 1) a stand-alone seasonal vaccination strategy reflecting current use of separate influenza and COVID-19 vaccines (“Stand-alone” strategy), and 2) a combination seasonal vaccination strategy incorporating mRNA-1083, with partial replacement of stand-alone influenza and COVID-19 vaccines (“Combination” strategy). Key outcomes included symptomatic infections, hospitalizations, deaths, healthcare system costs, societal costs, and the economically justifiable price (EJP) of mRNA-1083 within the Combination strategy using the commonly used Canadian willingness-to-pay (WTP) threshold of $50,000 per quality-adjusted life-year (QALY) gained [42–46].

## Methods

This study aligns with the Consolidated Health Economic Evaluation Reporting Standards checklist (Appendix Section 8) [47].

### Model Overview

To assess the impact of introducing a combination vaccine strategy in Canada, a previously published COVID-19 model was adapted and expanded to include influenza [48]. The original model was a static, two-state Markov model with a monthly cycle length, developed to predict COVID-19 infections over a one-year time horizon (September to August) [48]. COVID-19 and its consequences were modelled as an embedded decision tree to capture acute infection pathways within each cycle. For the present study, the model was modified with the addition of a separate decision tree representing acute influenza infection pathways. Although there is a risk of co-infection with both SARS-CoV-2 and influenza, Canadian data suggest that the prevalence of co-infection remains low (approximately 4.5%) [49]; therefore, the two infections were assumed to occur independently, with outcomes for each of the two infection types tracked separately.

All individuals enter the model in the Well health state at the start of the simulation (September, month 1). In the base-case, vaccine coverage for stand-alone influenza and COVID-19 vaccines is assumed to begin in September (with proportions for each consistent with observed coverage data) [23,48,50,51]. Within each cycle, individuals transition through influenza or COVID-19 decision tree pathways and either return to the Well state or transition to the Dead state following infection-related mortality. The states are depicted as coloured boxes at the start/end of the corresponding decision trees in Figures 1 and 2.

**Figure 1.**
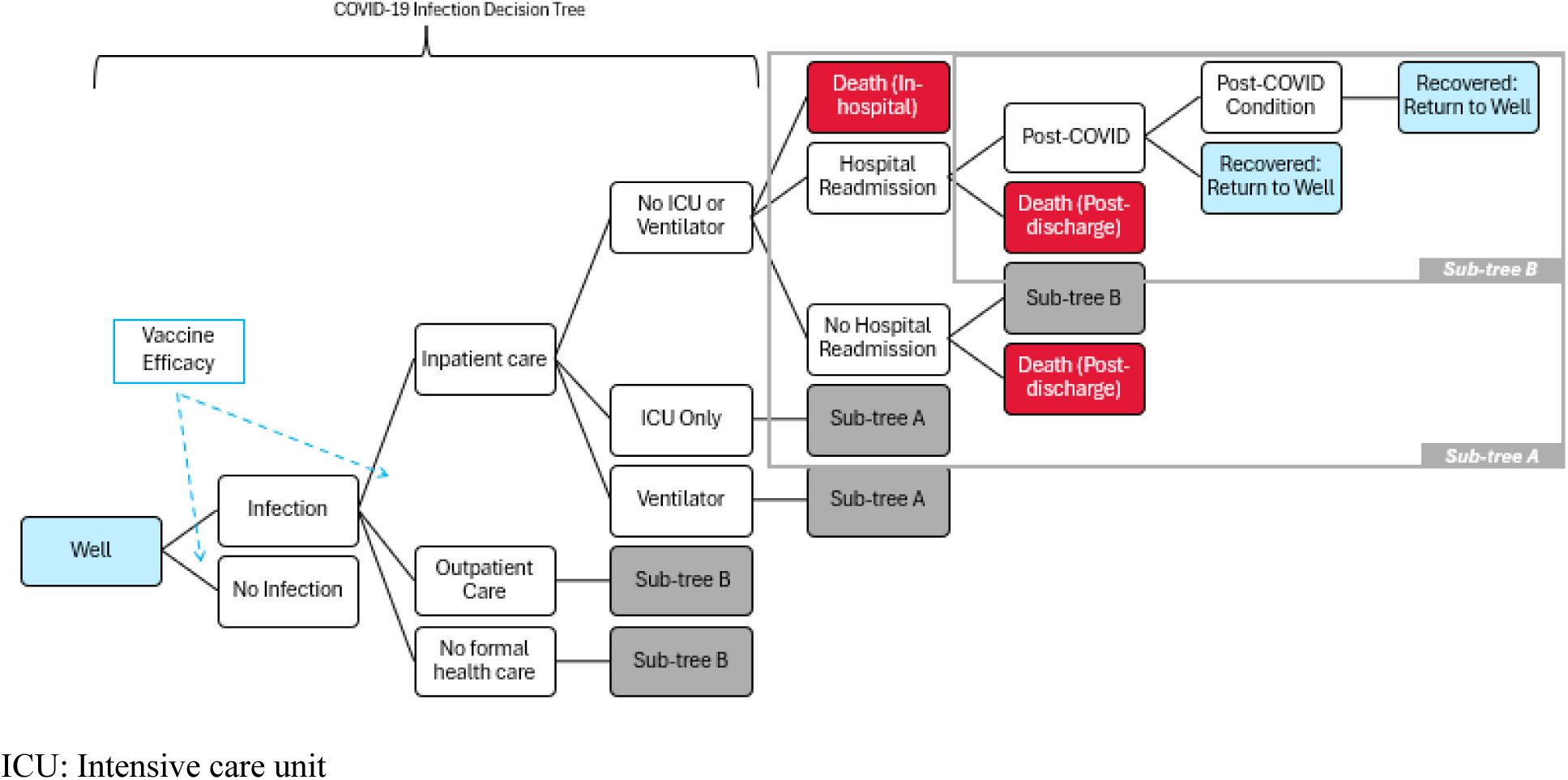
Overview of the COVID-19 infection decision tree within the Markov model. The blue squares represent the “Well” health state and the red squares represent the “Death” health state. Outpatient care is assumed to consist of emergency department and physician visits. Vaccine efficacy is represented by the dashed lines, while the infection pathways are represented by solid lines.

**Figure 2.**
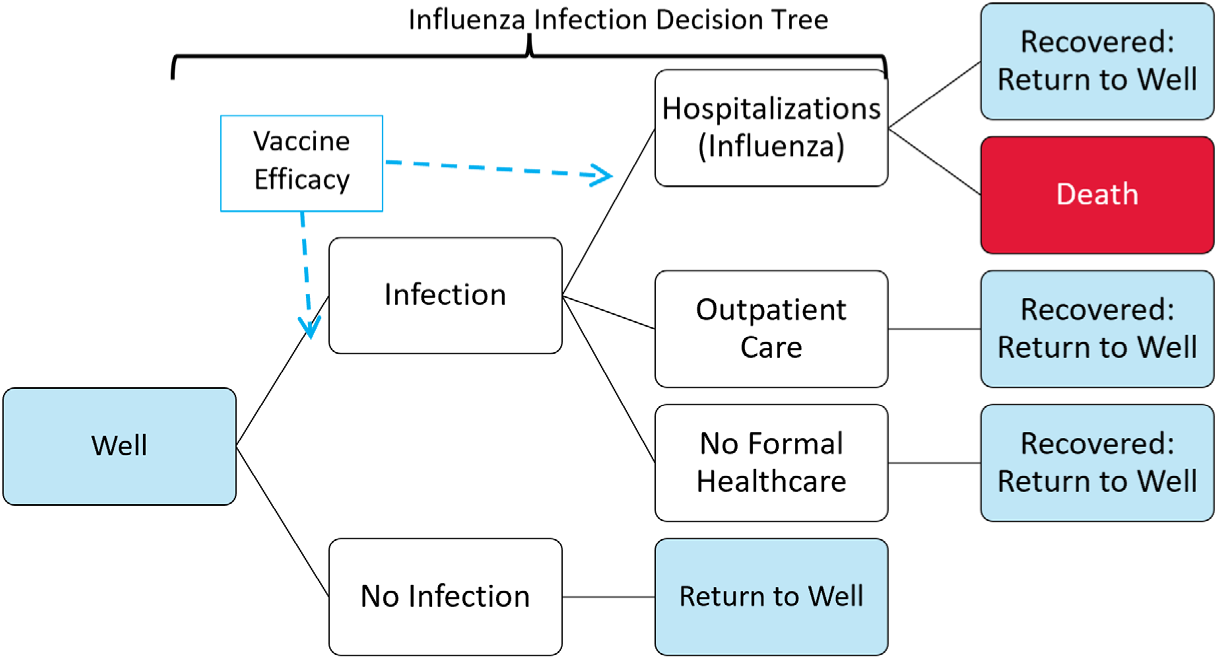
Overview of the influenza infection decision tree within the Markov model. The blue squares represent the “Well” health state and the red squares represent the “Death” health state. Outpatient care is assumed to consist of emergency department and physician visits. Vaccine efficacy is represented by the dashed lines, while the infection pathways are represented by solid lines.

Within the COVID-19 portion of the model (Figure 1), individuals in the Well state face a risk of SARS-CoV-2 infection, which can be reduced by vaccination. [48] Vaccination may further reduce the risk of hospitalization following infection. If an infection develops, individuals may not receive formal medical care, be treated on an outpatient basis, or be hospitalized.

COVID-related mortality is assumed to apply to hospitalized patients only. Those who die from COVID-19 transition to the Death state, while survivors return to the Well state [48].

In the influenza component shown in Figure 2, those in the Well state face the risk of an influenza infection every cycle. As with the COVID-19 decision tree, vaccinated individuals have a reduced risk of infection and subsequent hospitalization. Individuals who do not develop an infection return to the Well state. Those who develop an influenza infection may not receive medical care, be treated on an outpatient basis (i.e., emergency room or physician visit) [52], or be hospitalized. Influenza-related mortality is assumed to occur exclusively among hospitalized patients, with individuals who die transitioning to the Dead state.

Although the decision trees for COVID-19 and influenza are largely similar, key distinctions reflect differences in disease pathways, treatment patterns, and data availability for each infection type [48,52–55]. For example, the COVID-19 decision tree includes additional stratification of hospitalized patients by level of care (i.e., no intensive care unit [ICU] or mechanical ventilation [MV], ICU only, ICU with MV) [48,53]. COVID-19 patients are subject to risk of readmission and post-COVID sequelae [53,56]. The decision tree structures for each infection type were designed for consistency with previously published models and data availability [48,52,53].

### Target Population

For the base-case analysis, the target population comprised adults aged 65 years and older across Canada. Estimates of the population size, by age group, were obtained from Statistics Canada (Technical Appendix – section 4) [57].

### Comparator vaccination strategies

This study compared two seasonal vaccination strategies: a “Stand-alone” strategy and a “Combination” strategy.

The Stand-alone strategy represented current practice, wherein influenza and COVID-19 vaccines were administered separately based on 2024-2025 vaccine coverage data (see next section). The model tracks COVID-19 and influenza vaccine coverage independently.

Theoretically, the Stand-alone strategy comprised multiple mutually exclusive groups of vaccine recipients and unvaccinated individuals, categorized as those who received: 1) both COVID-19 and influenza vaccines; 2) the influenza vaccine only; 3) the COVID-19 vaccine only; or, 4) neither vaccine.

Under the Combination strategy, a proportion of individuals who would have received stand-alone influenza and/or COVID-19 vaccines in the Stand-alone strategy were assumed to receive mRNA-1083 instead, a single vaccine providing coverage against both infections. The remaining vaccinated population received stand-alone influenza and/or COVID-19 vaccines. Those individuals who did not receive either the COVID-19 or influenza vaccine in the Stand-alone strategy were assumed to remain unvaccinated in the Combination strategy. The impact of switching from stand-alone vaccination to mRNA-1083 across recipient groups is depicted in Figure 3.

**Figure 3.**
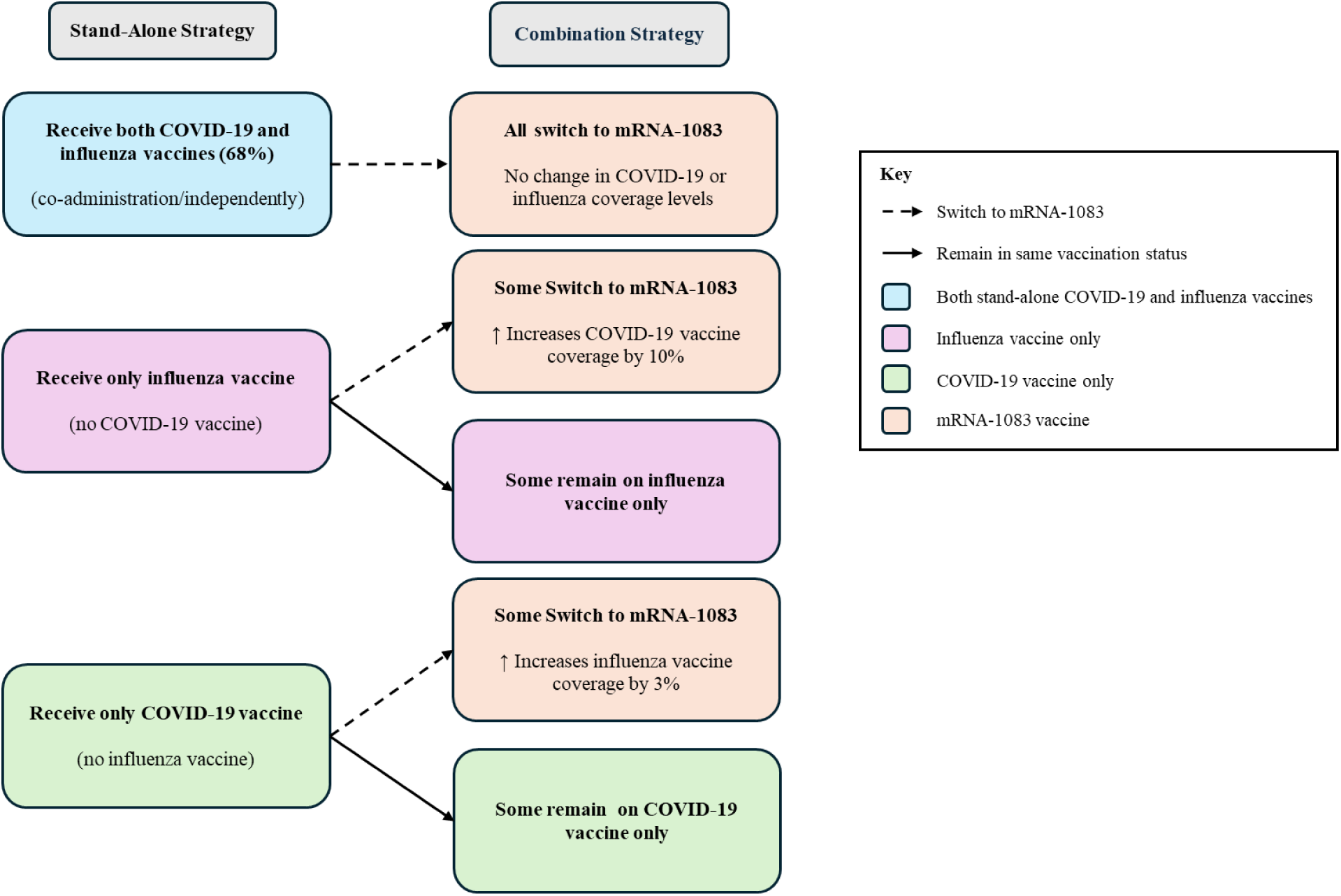
Impact of Switching from Stand-Alone Strategy to Combination Strategy on COVID-19 and Influenza Coverage Amongst Adults Aged 65 Years and Older Individuals who do not get either influenza or COVID-19 vaccine in the Stand-alone strategy remain unvaccinated in the Combination strategy. Based on PHAC data for the 2023-2024 season, 68.1% of adults aged ≥65 years who received both influenza and COVID-19 did so at the same visit [34].

Within each strategy, influenza vaccination was represented by IIV-Adj; this is a commonly used enhanced vaccine in Canada, given its equivalent clinical effectiveness to high-dose influenza vaccines and low price [14,58]. COVID-19 vaccination was represented by SPIKEVAX® (mRNA-1273; Moderna). Vaccine-specific uptake, efficacy, adverse events (AEs), and costs (acquisition, administration), were applied separately by vaccine type.

### Vaccine Coverage

The COVID-19 and influenza vaccine coverage inputs applied within this study were based on published Canadian data from the 2024-2025 season, consistent with the COVID-19 modeling study by Fust et al. (2025) [48], as more recent data are unavailable. The annual COVID-19 vaccine coverage for the 2024-2025 season for those aged ≥65 years (53.5%) was estimated from the Seasonal Respiratory Vaccination Coverage Survey from which vaccinations were assumed to begin in September [23,51]. Given that monthly estimates of vaccine coverage are required for the model, the distribution of vaccine coverage across months was obtained from PHAC COVID-19 data from the 2023-2024 season to replicate observed patterns of vaccination rollout (September), with peak coverage and completion reached by June (details provided in the Technical Appendix Section 2). Annual influenza vaccine coverage, by age group (63.2% for those aged ≥65 years), was estimated from the Seasonal Influenza Vaccination Coverage report, from which vaccination timing was assumed to begin in September [23]. Similar to the approach used for COVID-19 coverage, the distribution of vaccine coverage across months was obtained from PHAC influenza data [50]. Vaccination against influenza began in September, with peak coverage and completion reached by January. mRNA-1083 uptake was assumed to begin in September, with peak coverage and completion reached by January, consistent with influenza vaccination. Details are provided in the Technical Appendix (Section 2).

The Combination strategy required assumptions addressing the proportion of individuals receiving both stand-alone vaccines who switch to mRNA-1083. Based on PHAC data for the 2023-2024 season, 68.1% of adults aged ≥65 years who received both influenza and COVID-19 vaccines did so at the same visit [50]. Accordingly, this proportion was applied in the base-case to represent individuals switching from receipt of both stand-alone vaccines to mRNA-1083.

Additionally, the allocation of mRNA-1083 alongside the stand-alone vaccines within the Combination strategy was assumed to increase vaccine coverage. In the absence of Canadian evidence, US and European (i.e., France, Italy, and Germany) patient preference data examining the acceptability of an adult Combination vaccine for influenza and COVID-19 compared with stand-alone influenza and COVID-19 vaccines were applied to inform this effect [33,59,60]. A proportion of influenza-only vaccine recipients were assumed to switch to mRNA-1083, increasing COVID-19 coverage by 10% (63.5% versus Stand-alone strategy: 53.5%). The total vaccinations administered did not change. Rather, the proportion of individuals protected against COVID-19 increased via mRNA-1083 by extending protection to those who would otherwise have received influenza vaccination only. For influenza, an increase in vaccine coverage of 3% was assumed (66.2% versus Stand-alone strategy: 63.2%), occurring in the model as a result of people switching from receiving the stand-alone COVID-19 vaccine to mRNA-1083 [59,60].

### Infection Incidence

Data on the monthly incidence of symptomatic COVID-19 and influenza in the Canadian population in those who did not receive an annual vaccine is not available. Consistent with Fust et al. (2025) [48], monthly incidence for COVID-19 and influenza infections was therefore estimated using monthly age-specific hospitalization rates per 100,000 from the 2023-2024 and 2024-2025 seasons, based on Government of Canada data and Miranda et al. (2024) [3,53,61,62]. Vaccine coverage, effectiveness, and the probability of hospitalization, conditional on symptomatic infection, were incorporated in the estimation of incidence (per Fust et al. (2025) [48], see Technical Appendix Section 1). Base-case COVID-19 and influenza incidence estimates were derived by averaging modeled monthly incidence over the September 2023–August 2024 and September 2024–August 2025 periods.

### Vaccine Effectiveness against COVID-19: mRNA-1273 and mRNA-1083

Vaccine effectiveness (VE) and waning assumptions for mRNA-1273 [63] and mRNA-1083 [64] against COVID-19 outcomes were based on published studies. Initial VE for mRNA-1273 was derived from a retrospective observational cohort study evaluating protection against the KP.2 variant during the 2024–2025 season, using medically-attended COVID-19 as a proxy for infection and COVID-19-related hospitalizations [63]. Initial VE for mRNA-1083 was assumed to be equivalent to that of its COVID-19 vaccine component, i.e., mRNA-1283, with relative vaccine efficacy (rVE) estimates derived from the pivotal phase 3 clinical trial (NextCOVE) comparing mRNA-1283 with mRNA-1273 (13.5% against infection; 38.1% against hospitalization) [64]. Additional rVE estimation details are described in Fust et al. (2025) [48]. Consistent with the waning assumptions applied to mRNA-1283 in Fust et al. (2025)[48], VE for mRNA-1273 and mRNA-1083 was assumed to decline by 4.75% per month against infection [65] and 2.46% per month against hospitalization [66].

### Vaccine Effectiveness against influenza: IIV-Adj and mRNA-1083

The initial VE for IIV-Adj was calculated based on data from Separovic et al. (2025) [67], who reported interim 2024-2025 VE estimates from the Canadian Sentinel Practitioner Surveillance Network. The study estimated that the adjusted VE against laboratory-confirmed influenza for those aged ≥65 years was 59% from late October 2024 to mid-January 2025. As this represents an average VE over 3 months, the initial VE value for month 1 (67% for those aged ≥65 years) was back-calculated using a waning rate of 8% [68]. The initial VE against hospitalization was assumed to be equal to VE against infection (i.e. no incremental protection). In the model, IIV-Adj VE was assumed to wane by 8% per month [68].

Immunogenicity data from the mRNA-1083-P301 clinical trial comparing mRNA-1083 to high-dose quadrivalent inactivated influenza vaccine demonstrate that for the 3 clinically relevant circulating strains of influenza, mRNA-1083 is associated with a higher immune response than enhanced dose vaccine[34]. Accordingly, for the base-case, the VE assumptions modeled for mRNA-1083 against influenza were equal to the initial VE and waning rate applied for IIV-Adj (i.e., no additional protection against influenza for mRNA-1083 is modeled).

### Probabilities for the COVID-19 and Influenza Infection Consequences Decision Trees

COVID-19 and influenza estimates, along with ranges used in deterministic sensitivity analyses, are provided in Table 3. Estimates for COVID-19 are consistent with those provided in Fust et al. (2025) [48].

**Table 2.**
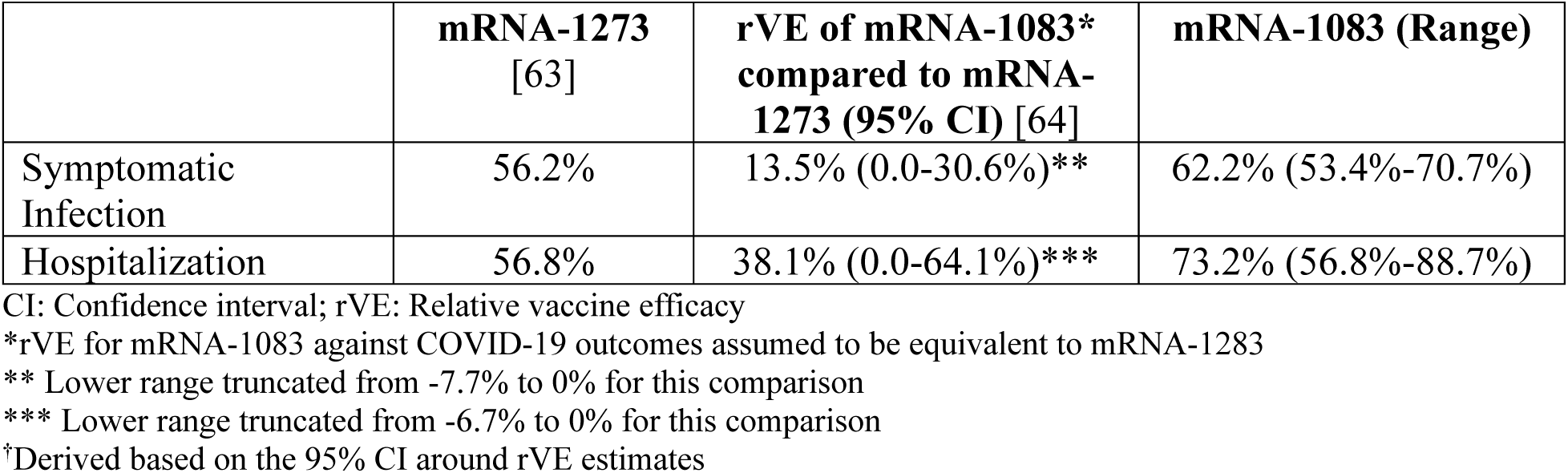
Initial vaccine effectiveness values (%) (COVID-19)

**Table 3.** Key decision tree probabilities for COVID-19 and influenza, base-case and range for deterministic sensitivity analyses.

| Parameter | Base (Range) |  |  | Source |
| --- | --- | --- | --- | --- |
| Distribution of Care Following a Symptomatic Infection |  |  |  |  |
|  | % Hospitalized | % No Formal Care | % Outpatient Care |  |
| COVID-19 | 3.9% (±10%)* | 58.6% | 37.5% | Derived based on UK data [69];US CDC Influenza Data [70] |
| Influenza<sup>†</sup> | 9.1% (±10%) | 34.9% | 56.0% | US CDC Burden of Illness Data [71] |
| In-Hospital Distribution of Care (COVID-19 Only) |  |  |  |  |
|  | No ICU or Ventilator | ICU Only | ICU with Ventilator |  |
| COVID-19 | 86.8% | 8.1% | 5.1% | CIHI [72] |
| In-Hospital Mortality<sup>‡</sup> (%) |  |  |  |  |
| COVID-19 | 15.6% (±25%) |  |  | Miranda 2024 [53] |
| Influenza | 19.0% (±25%) |  |  | Fisman et al. (2011) [52] |
| Post-Discharge Mortality (%; COVID-19 Only)<sup>‡</sup> |  |  |  |  |
| COVID-19 | 3.7% |  |  | Qian 2025 [56]<sup>§</sup> |
| Post-Infection (%; COVID-19 Only)<sup>§§</sup> |  |  |  |  |
| COVID-19 | 100% |  |  | Fust 2025 [48] |
| Post- COVID Condition (%; COVID-19 Only)<sup>**</sup> |  |  |  |  |
| COVID-19 | 8.1% |  |  | US CDC 2025 [73] |

CDC: Centers for Disease Control and Prevention; CIHI: Canadian Institute for Health Information; ICU: Intensive Care Unit; UK: United Kingdom; US: United States
\*Hospitalized COVID-19 patients are subject to risk of readmission; however, this probability is assumed to be 0% to avoid double-counting with the hospitalization cost estimates (which already reflect readmission)
<sup>†</sup>Due to a lack of data specific to the Canadian setting, the age-specific probabilities of hospitalization, outpatient visits, and no formal care for the general population were estimated from the US Centers for Disease Control and Prevention (CDC) [46] preliminary estimated influenza disease burden for the 2023-2024 season (2024-2025 estimates were not available at the time this study was conducted).
<sup>‡</sup>Only hospitalized patients are subject to risk of death from COVID-19 or influenza
<sup>§</sup>Calculated by subtracting the reported in-hospital mortality from the 60-day mortality
<sup>§§</sup>Post-infection QALY tolls are assigned to all patients (hospitalized or outpatient) who survive the acute phase of COVID-19 infection.
<sup>\*\*</sup>Post-COVID Condition (Long COVID) is associated with lost productivity-related costs only and is therefore only reflected in analyses performed from the societal perspective.

### Adverse Event Rates

Grade 3 and 4 AE rates for each vaccine (stand-alone influenza, stand-alone COVID-19, mRNA-1083) were estimated from the mRNA-1083 clinical trial [34]. Solicited local and systemic AEs were assessed through 7 days after vaccination, while unsolicited AEs and severe AEs were assessed up to 28 days following vaccination[34]. Serious AEs, medically attended AEs, AEs of special interest, and AEs leading to discontinuation were assessed through the end of the clinical trial (up to 181 days)[34]. Details are presented in the Technical Appendix (Section 5).

### Quality of Life

The QALY loss estimates associated with infections, vaccine-related AEs, and mortality were derived from published sources (Table 4). Lifetime QALY losses associated with premature death due to COVID-19 and influenza infection during the one-year time horizon, discounted to present value at an annual rate of 1.5%, were also included [74].

**Table 4.** QALY Loss Inputs.

| Parameter | QALY loss (Range) | Source |
| --- | --- | --- |
| **Adverse Events** |  |  |
| Grade 3 (Local or Systemic) | 0.0004 | Walter 2024 [75] |
| Grade 4 Systemic | 0.0027 | Miranda 2024 [76] |
| Anaphylaxis | 0.00192 | Prosser 2019 [77] |
| Myocarditis/Pericarditis | 0.00064 |  |
| Infection-Related Myocarditis | 0.00192 |  |
| Guillain-Barré Syndrome | 0.0227 | Calculated based on DeLuca et al. (2023) [78] |
| **COVID-19 infection-related** |  |  |
| No formal care | 0.0046 (0.0018 - 0.0074) | UM-CDC [79] |
| Outpatient care | 0.0046 (0.0018 - 0.0074) |  |
| Hospitalization: |  |  |
| No ICU or Ventilator | 0.0174 (0.0038 - 0.0310) | UM-CDC [79] |
| ICU Only | 0.0394 (0.0231 – 0.0583) |  |
| ICU with Ventilator | 0.0394 (0.0231 – 0.0583) |  |
| Post-acute care: |  |  |
| Outpatient care | 0.023 | Sandmann 2022 [80] |
| Hospitalized | 0.103 | PHOSP 2022 [81] |
| **Influenza infection-related\*** |  |  |
| No formal care | 0.029 (0.023 – 0.035) | Fisman DN et al. 2011 [52] |
| Outpatient care | 0.029 (0.023 – 0.035) |  |
| Hospitalization | 0.029 (0.023 – 0.035) |  |

ICU: Intensive care unit; QALY: Quality-adjusted life-year
\*Fisman et al. [52] provides age-specific estimates; however, they are not provided by location of care. Accordingly, the same QALY losses were assigned to those with no formal care, outpatient care, and hospitalization.

### Healthcare Costs

Base-case cost estimates were selected to reflect the publicly funded health system perspective [82] and are reported in 2025 Canadian dollars. As the model time horizon was one year, costs were not discounted [83]. Cost inputs were predominantly obtained from published Canadian literature and data sources. The inputs included the cost of vaccine acquisition, vaccine administration, management of vaccine-related AE, and infection-related healthcare costs by care setting (Table 5).

**Table 5.**
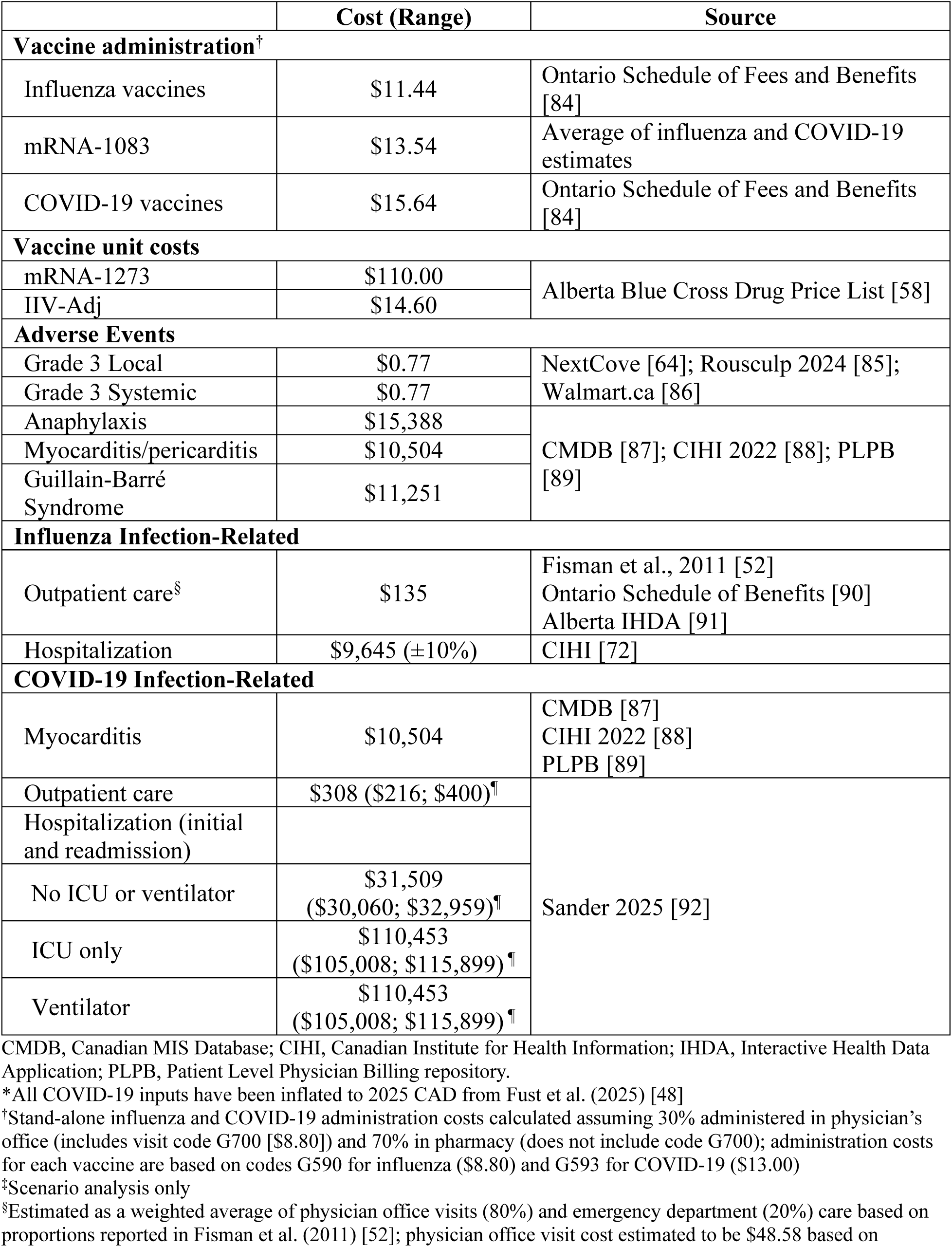
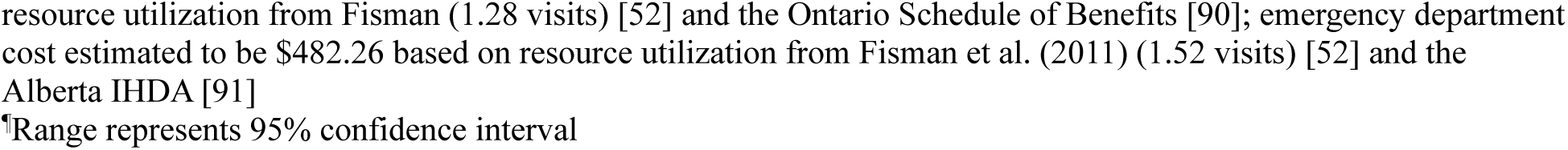
Healthcare Cost Inputs*.

|  | Cost (Range) | Source |
| --- | --- | --- |
| Vaccine administration† |  |  |
| Influenza vaccines | \$11.44 | Ontario Schedule of Fees and Benefits [84] |
| mRNA-1083 | \$13.54 | Average of influenza and COVID-19 estimates |
| COVID-19 vaccines | \$15.64 | Ontario Schedule of Fees and Benefits [84] |
| Vaccine unit costs |  |  |
| mRNA-1273 | \$110.00 | Alberta Blue Cross Drug Price List [58] |
| IIV-Adj | \$14.60 | |
| Adverse Events |  |  |
| Grade 3 Local | \$0.77 | NextCove [64]; Rousculp 2024 [85]; Walmart.ca [86] |
| Grade 3 Systemic | \$0.77 | |
| Anaphylaxis | \$15,388 | CMDB [87]; CIHI 2022 [88]; PLPB [89] |
| Myocarditis/pericarditis | \$10,504 | |
| Guillain-Barré Syndrome | \$11,251 | |
| Influenza Infection-Related |  |  |
| Outpatient care§ | \$135 | Fisman et al., 2011 [52]<br>Ontario Schedule of Benefits [90]<br>Alberta IHDA [91] |
| Hospitalization | \$9,645 (±10%) | CIHI [72] |
| COVID-19 Infection-Related |  |  |
| Myocarditis | \$10,504 | CMDB [87]<br>CIHI 2022 [88]<br>PLPB [89] |
| Outpatient care | \$308 (\$216; \$400)¶ | Sander 2025 [92] |
| Hospitalization (initial and readmission) |  |  |
| No ICU or ventilator | \$31,509<br>(\$30,060; \$32,959)¶ | |
| ICU only | \$110,453<br>(\$105,008; \$115,899)¶ | |
| Ventilator | \$110,453<br>(\$105,008; \$115,899)¶ | |

CMDB, Canadian MIS Database; CIHI, Canadian Institute for Health Information; IHDA, Interactive Health Data Application; PLPB, Patient Level Physician Billing repository.
\*All COVID-19 inputs have been inflated to 2025 CAD from Fust et al. (2025) [48]
<sup>†</sup>Stand-alone influenza and COVID-19 administration costs calculated assuming 30% administered in physician's office (includes visit code G700 [\$8.80]) and 70% in pharmacy (does not include code G700); administration costs for each vaccine are based on codes G590 for influenza (\$8.80) and G593 for COVID-19 (\$13.00)
<sup>‡</sup>Scenario analysis only
<sup>§</sup>Estimated as a weighted average of physician office visits (80%) and emergency department (20%) care based on proportions reported in Fisman et al. (2011) [52]; physician office visit cost estimated to be \$48.58 based on
resource utilization from Fisman (1.28 visits) [52] and the Ontario Schedule of Benefits [90]; emergency department cost estimated to be \$482.26 based on resource utilization from Fisman et al. (2011) (1.52 visits) [52] and the Alberta IHDA [91]
\*Range represents 95% confidence interval

### Scenario Analyses: Vaccine Effectiveness and Coverage

Scenario analyses were conducted to assess the impact of VE and coverage, both independently and jointly, on clinical outcomes and economic results (Table 6). For several of the Combination strategy scenarios, COVID-19 vaccine coverage increased by to 15% (base-case: 10%) to evaluate the impact of improved uptake, while a scenario assuming no increase in COVID-19 coverage (0%) was examined to isolate the effects of improved VE with mRNA-1083 and potential administration fee savings. Additional scenarios included the assumption that mRNA-1083 had no rVE benefit compared to mRNA-1273 to further isolate the impact of increased COVID-19 coverage. Finally, scenarios incorporating an influenza VE benefit for mRNA-1083, which is not subject to egg-adaptation, were explored. Stein et al conducted an analysis comparing a cell-based (IIV-cc) and an egg-based (IIV-SD) influenza vaccine across 3 influenza seasons and estimated the rVE of IIV-cc compared to IIV-SD to be 12.4% [41]. We conservatively assumed that the rVE mRNA-1083 compared to IIV-Adj would be 50% of that value, or 6.2%.

**Table 6.** Vaccine Effectiveness and Coverage Scenario Analyses.

|  | **Rationale** | **rVE against COVID-19** | **rVE against Influenza** | **COVID-19 Coverage Increase** | **Influenza Coverage Increase** |
| --- | --- | --- | --- | --- | --- |
| **Base-Case** | Estimate the impact of improved VE with mRNA-1083 against COVID-19 (compared to mRNA-1273) and increases in COVID-19 and flu VCRs | rVE = 13.5% (Infection)<br>rVE = 38.1% (Hospitalization) | rVE = 0% (Infection)<br>rVE = 0% (Hospitalization) | **10%** | **3%** |
| **Scenarios** |  |  |  |  |  |
| **Increased COVID-19 VCR** | Demonstrate impact of increasing COVID-19 VCR further (from 10% in base-case) | Base-Case | Base-Case | 15% | Base-Case |
| **No increase in COVID-19 or influenza VCR** | Isolates the impact of improved VE against COVID-19 with mRNA-1083 | Base-Case | Base-Case | 0% | 0% |
| **No rVE against COVID-19** | Isolates the impact of increasing COVID-19 and influenza VCR | rVE=0% (Infection)<br>rVE=0% (Hospitalization) | Base-Case | Base-Case | Base-Case |
| **No rVE against COVID-19, increased COVID-19 VCR** | Isolates the impact of increasing COVID-19 VCR further | rVE=0% (Infection)<br>rVE=0% (Hospitalization) | Base-Case | 15% | Base-Case |
| **No rVE against COVID-19, no increase in COVID-19 or influenza VCR** | Isolates the impact of administration fee savings with mRNA-1083 | rVE=0% (Infection)<br>rVE=0% (Hospitalization) | Base-Case | 0% | 0% |
| **Include increased rVE against influenza** | Combined impact of improved VE with mRNA-1083 for both COVID & influenza and increasing COVID-19 and Influenza VCRs | Base-Case | rVE = 6.2% (Infection)<br>rVE = 6.2% (Hospitalization) | Base-Case | Base-Case |
| **Include increased rVE against influenza, increased COVID-19 VCR** | Combined impact of improved VE with mRNA-1083 for both COVID & influenza and increasing COVID-19 and Influenza VCRs | Base-Case | rVE = 6.2% (Infection)<br>rVE = 6.2% (Hospitalization) | 15% | Base-Case |
| **Include increased rVE against influenza, no increase in COVID-19 or influenza VCR** | Isolates the impact of improved VE with mRNA-1083 for both COVID & influenza | Base-Case | rVE = 6.2% (Infection)<br>rVE = 6.2% (Hospitalization) | 0% | 0% |

rVE: Relative vaccine effectiveness; VCR: Vaccine coverage rate; VE: Vaccine effectiveness

### Scenario Analyses: Administration Fees

The distribution of COVID-19 and influenza vaccine administrations across care settings (pharmacy versus a physician’s office) in Canada is not well characterized. As such, in the base-case, a 30% physician and 70% pharmacy split was assumed. Given the uncertainty, various scenario analyses related to the administration costs for stand-alone COVID-19 vaccines, stand-alone influenza vaccines, and mRNA-1083 were performed, including a scenario in which mRNA-1083 was assigned an administration fee premium equivalent to 2.5 times the base-case value (Table 7).

**Table 7.** Vaccine administration costs (base-case and scenario analyses)

|  | **mRNA-1083\*** | **mRNA-1273** | **Influenza (IIV-Adj)** | **Administration Assumptions [93-95]** |
| --- | --- | --- | --- | --- |
| Base-Case | \$13.54 | \$15.64 | \$11.44 | 30% physician's office and 70% pharmacy for COVID-19 and influenza<sup>†</sup> |
| Pharmacy Administration Only<sup>‡</sup> | \$10.90 | \$13.00 | \$8.80 | 100% pharmacy administration for both COVID-19 & Influenza |
| Physician Office Administration Only | \$19.70 | \$21.80 | \$17.60 | 100% physician office administration for both COVID-19 & Influenza |
| Increased mRNA-1083 Administration Cost | \$33.85 | \$15.64 | \$11.44 | Administration cost for mRNA-1083 is 2.5x the base-case |

\*mRNA-1083 administration fees calculated as the average of mRNA-1273 & IIV-Adj fees, with the exception of the "Increased mRNA-1083 Administration Cost" scenario
<sup>†</sup>Vaccines administered in a physician's office include a visit fee of \$8.80 (Code G700) in addition to the administration fees (\$13.00 for COVID-19 vaccines, \$8.80 for influenza vaccines).
<sup>‡</sup>This scenario is equivalent to base case but assuming that any vaccine being administered within a physician's office is at another scheduled visit so that the physician's office visit fee is set to 0.

### Scenario Analyses: Other

Several scenario analyses were conducted to address the impact of alternate vaccine coverage rates, VE, comparator selection, and cost perspective. In a VE scenario, the rVE of mRNA-1083 against COVID-19 hospitalization was equivalent to the rVE for infection (i.e., no additional protection against hospitalization). Multiple comparator scenarios assessed the use of Comirnaty® (BNT-162b2; Pfizer-BioNTech) in place of mRNA-1273 and IIV-HD in place of IIV-Adj. In these analyses, VE for BNT-162b2 was estimated as described by Fust et al. (2025) (Technical Appendix Section 3) [48], with AE rates presented in the Technical Appendix. For IIV-HD, VE and AE rates were assumed equivalent to IIV-Adj based on NACI guidance on influenza vaccination for adults aged ≥65 years [14]. The unit prices of BNT162b2 and IIV-HD were $107.65 and $67.36, respectively, as obtained from Alberta’s public drug formulary [58]. Finally, a scenario analysis was conducted from the societal perspective [82], which includes time loss from work associated with vaccine administration, AEs, and COVID-19 or influenza-related infections (Technical Appendix Section 6).

### Deterministic Sensitivity Analyses

Deterministic sensitivity analyses were undertaken to evaluate the impact of key parameters on the Combination strategy’s EJP. mRNA-1083 VE was varied using the 95% confidence intervals (CI) of rVE estimates for mRNA-1083 versus mRNA-1273 (Table 2). Waning rates against infection were varied from 3.05% to 6.75% per month, while that against hospitalization varied from 1.37% to 3.87% per month [65,96]. COVID-19 and influenza incidence were varied using season-specific estimates from 2023-2024 and 2024-2025 independently. In addition, COVID-19-and influenza-related hospitalization mortality, costs, and QALY losses were also varied as indicated in Tables 3-5. Where available, model parameters were varied according to 95% CIs or reported ranges. For all other DSAs, parameters were varied by ±10% or ±25% of their base-case value.

## Results

### Base-case

The base-case clinical results are presented in Table 8. The increase in COVID-19 vaccination coverage and the higher VE of mRNA-1083 compared to mRNA-1273 with the Combination strategy leads to a reduction in the number of COVID-19-related symptomatic infections (71,074 [6%]), hospitalizations (5,008 [12%]), and deaths (935 [12%]) (Table 8). Although mRNA-1083 is considered to have equal VE to IIV-Adj in the base-case, the Combination strategy leads to an approximate 2% reduction in the number of influenza symptomatic infections, hospitalizations, and deaths due to the increase in influenza coverage. With the Combination strategy, there are more local and systemic adverse events, but fewer cases of anaphylaxis and Guillain-Barré Syndrome (GBS) (Appendix Table 12).

**Table 8.** Base-Case Results: Clinical Outcomes.

| Outcome | Stand-Alone Strategy |  | Combination Strategy |  | Difference<br>(Combination Strategy<br>– Stand-Alone<br>Strategy) |  | % Change |  |
| --- | --- | --- | --- | --- | --- | --- | --- | --- |
|  | COVID-19 | Influenza | COVID-19 | Influenza | COVID-19 | Influenza * | COVID-19 | Influenza |
| Symptomatic infections | 1,124,836 | 257,067 | 1,053,762 | 253,082 | -71,074 | -3,985 | -6% | -2% |
| Outpatient cases | 423,412 | 143,958 | 399,012 | 141,726 | -24,400 | -2,232 | -6% | -2% |
| Hospitalizations** | 42,013 | 23,370 | 37,005 | 23,007 | -5,008 | -362 | -12% | -2% |
| Post-COVID Condition | 36,912 | -- | 34,615 | -- | -2,297 | -- | -6% | -- |
| Deaths | 7,841 | 4,440 | 6,907 | 4,371 | -935 | -69 | -12% | -2% |

\* Influenza outcomes do not significantly change as mRNA-1083 is considered to have equal VE to IIV-Adj in the base case analysis; slight differences in influenza outcomes may be observed due to the change in the number of deaths in the model with the Combination strategy and the 3% increase in influenza uptake
\*\*COVID-19 and influenza-related hospitalizations

The detailed costs associated with the Stand-Alone and Combination strategies are shown in Table 9. Overall, the costs of vaccine administration decrease with the Combination strategy (31% reduction) because of the decline in the number of injections occurring as a result of individuals switching from receiving both the COVID-19 and influenza vaccines in the Stand-Alone strategy to mRNA-1083. The overall cost of adverse events is also lower (10% reduction), driven primarily by a decrease in severe AEs. Finally, the decrease in COVID-19 and influenza outcomes in the Combination strategy leads to a decrease in the expected COVID-19 and influenza-related treatment costs, which are reduced by 11% and 2%, respectively. A detailed table presenting QALYs lost with each strategy is presented in the Technical Appendix (Table 11). Although there is an increase in QALYs lost due to AEs in the Combination strategy compared to the Stand-alone strategy, overall QALYs gained increased with the Combination strategy due to reductions in COVID-19 and influenza clinical outcomes. The base-case economically justifiable cost of the mRNA-1083 strategy is $304.11.

**Table 9.** Base-Case Results: Costs (Healthcare Perspective)

| Cost Category | Stand-Alone Strategy |  | Combination Strategy |  | Difference (%) (Combination – Stand-Alone Strategy)* |  |
| --- | --- | --- | --- | --- | --- | --- |
|  | COVID-19 | Influenza | COVID-19 | Influenza | COVID-19 | Influenza |
| Outpatient care | \$130,551,722 | \$19,479,445 | \$123,028,266 | \$19,177,468 | -\$7,523,456<br>(-6%) | -\$301,977<br>(-2%) |
| Hospitalizations | \$1,760,010,876 | \$225,395,759 | \$1,550,213,895 | \$221,901,592 | -\$209,796,981<br>(-12%) | -\$3,494,167<br>(-2%) |
| COVID-19 infection-related myocarditis | \$21,422,678 | -- | \$20,069,059 | -- | -\$1,353,619<br>(-6%) | -- |
| **Total infection-related costs** | **\$1,911,985,276** | **\$244,875,204** | **\$1,693,311,220** | **\$241,079,059** | **-\$218,674,056**<br>**(-11%)** | **-\$3,796,145**<br>**(-2%)** |
| Vaccination administration costs | \$130,477,599 | | \$90,440,471 | | -\$40,037,128 (-31%) | |
| Vaccine-related adverse events | \$977,432 | | \$878,697 | | -\$98,735 (-10%) | |
| **Total vaccination-related costs** | **\$131,455,031** | | **\$91,319,168** | | **-\$40,135,863 (-31%)** | |

\*Deaths in the model are removed each month; accordingly, the denominator changes. When the coverage of COVID-19 vaccination is increased in the Combination Strategy, the denominator changes as fewer COVID-19 related deaths occur. The small differences in the influenza strategy costs are driven by this and by the increase in influenza vaccination coverage.

**Table 10.** Economically Justifiable Price: Base-Case, Coverage, and Vaccine Effectiveness Scenarios.

|  | Coverage Scenarios |  |  |  |  |  |
| --- | --- | --- | --- | --- | --- | --- |
| Vaccine Effectiveness | Base-Case |  | Increased COVID-19 VCR |  | No Increase in COVID-19 & Influenza VCR |  |
| Number of Hospitalizations Averted |  |  |  |  |  |  |
|  | COVID-19 | Influenza | COVID-19 | Influenza | COVID-19 | Influenza |
| Base-Case | 5,008 | 362 | 6,237 | 362 | 2,334 | -1 |
| No rVE against COVID-19 | 1,851 | 363 | 2,776 | 363 | 0 | 0 |
| Include increased rVEs against influenza (due to egg adaptation) | 5,008 | 677 | 6,237 | 708 | 2,334 | 231 |
| Number of Deaths Averted |  |  |  |  |  |  |
|  | COVID-19 | Influenza | COVID-19 | Influenza | COVID-19 | Influenza |
| Base-Case | 935 | 69 | 1,164 | 69 | 436 | 0 |
| No rVE against COVID-19 | 345 | 69 | 518 | 69 | 0 | 0 |
| Include increased rVEs against influenza (due to egg adaptation) | 935 | 129 | 1,164 | 135 | 436 | 44 |
| Economically Justifiable Price |  |  |  |  |  |  |
| Base-Case | \$304.11 | | \$319.19 | | \$249.93 | |
| No rVE against COVID-19 | \$191.65 | | \$207.26 | | \$137.15 | |
| Include increased rVEs against influenza (due to egg adaptation) | \$313.40 | | \$328.48 | | \$259.22 | |

rVE: Relative vaccine effectiveness; VCR: Vaccine coverage rate

**Table 11.**
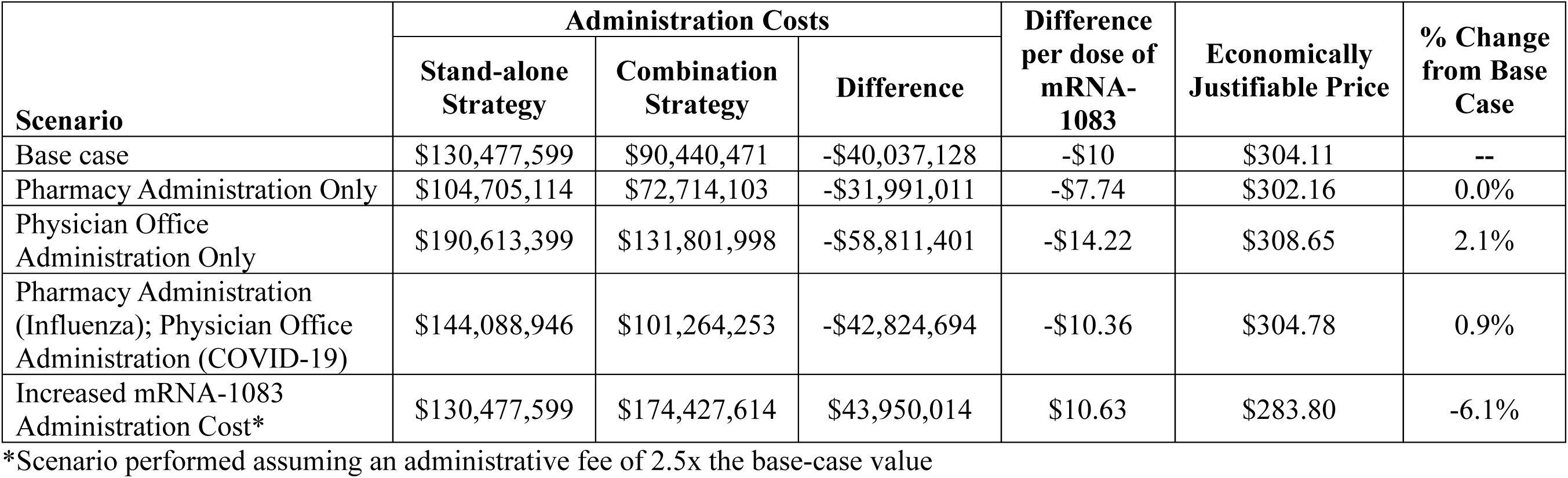
Administration Cost Scenario Results.

### Coverage Scenarios Results

Results of the scenarios examining the impact of vaccine coverage rates and VE, in isolation and in combination, on the hospitalizations averted, deaths averted and the EJP are presented in Table 10 below. Additional clinical results and the numbers of vaccine doses administered for these scenarios are presented in the Technical Appendix (Tables 13 and 15).

Two scenarios were performed varying only the VE assumptions. When mRNA-1083 has the same protection against COVID-19 as mRNA-1273 (i.e., equivalent VE against COVID-19), there are fewer COVID-19 hospitalizations averted by the Combination strategy (1,851 compared to 5,008for the base-case) and the EJP falls to $191.65. When mRNA-1083 has greater VE than IIV-Adj because IIV-Adj is impacted by egg adaptation, there are more influenza-related hospitalizations prevented by the Combination strategy as compared to the base-case (677 vs. 362 respectively); and the EJP increases to $313.40.

Three scenarios were performed utilizing an increase in the COVID-19 VCR of 15% (instead of the base-case value of 10%) with varying VE assumptions. If the Combination strategy were to increase COVID-19 VCR by 15%, there are more COVID-19 hospitalizations averted by the Combination strategy (6,237 compared to 5,008 for the base-case) and correspondingly, the EJP would increase to $319.19. Assuming that mRNA-1083 has no additional protection against COVID-19 (i.e., no rVE against COVID-19), with the 15% increase in the COVID-19 VCR, the EJP falls to $207.26. In this scenario, there are fewer COVID-19 hospitalizations averted (2,776) compared to the base-case (5,008). Including mRNA-1083 VE benefits for both COVID-19 and influenza (due to egg adaptation) and a 15% increase in COVID-19 VCR, leads to more COVID-19 and influenza related hospitalizations averted by the Combination strategy (6,237 and 708 compared to 5,008 and 362 in the base-case, respectively); accordingly, this scenario increases the EJP to $328.48.

Three additional scenarios were performed assuming that mRNA-1083 would not increase either COVID-19 or influenza VCRs within the Combination strategy. In a scenario designed to isolate the impact of the base-case VE assumptions (i.e., mRNA-1083 offers additional protection against COVID-19 as compared to mRNA-1273), the EJP is $249.93, due to reductions in COVID-19 and influenza hospitalizations averted by the Combination strategy as compared to the base-case (2,334 and 1 compared to 5,008 and 362, respectively). If no rVE against COVID-19 for mRNA-1083 is assumed, the EJP falls to $137.15; this estimate represents the impact of administration fee savings with mRNA-1083 occurring as a result of individuals switching from both stand-alone vaccines to the single mRNA-1083 vaccine. This EJP is still higher than the sum of the unit cost of the two single vaccines (mRNA-1273 + Fluad = $124.60) because of the cost savings associated with the administration of fewer injections. Finally, in a scenario including mRNA-1083 protection against influenza due to egg adaptation (in addition to COVID-19), the EJP decreases to $259.22, representing the impact of improved VE with mRNA-1083 against both COVID-19 and influenza. In this scenario, COVID-19 hospitalizations averted by the Combination strategy fall to 2,334 while influenza-related hospitalizations averted fall to 231.

### Additional Scenario and Sensitivity Analyses Results

Results of the various administration cost scenarios designed to assess the uncertainty in vaccine administration fees are presented in Table 11 below. Varying the administration fee for a combination vaccine such as mRNA-1083 cause the EJP to range from $283.80 to $308.65.

Increasing the administration fee for mRNA-1083 to 2.5 times the base-case value yields an EJP of $283.80, a 7% reduction from the base-case EJP. Administration costs for the Stand-alone strategy were $130,477,599 compared to $174,427,614 for the Combination strategy, leading to an increase in administration costs of $43,950,014 for the Combination strategy. Assuming all vaccines are administered in the physician’s office increases the EJP to $308.65 (1.5% increase), yielding administration fee savings of $58,811,401 for the Combination strategy (total administration costs of $131,801,998) compared to the Stand-alone strategy (total administration costs of $190,613,399). The administration costs saved for each mRNA-1083 vaccine administered ranged from $7.74 to $14.22 per dose of mRNA-1083 administered, while the scenario assuming an increased administration fee for mRNA-1083 increased administration costs to $10.63 per dose. All other administration cost scenarios led to changes in the EJP of <1%.

Tables summarizing the impact of all remaining scenarios and sensitivity analyses on clinical outcomes (symptomatic infections, hospitalizations, and deaths) and on the EJP are presented in the Technical Appendix (Tables 13 and 14). The scenario with the largest impact on the EJP is when the mRNA-1083 rVE for COVID-19 hospitalization is set equal to infection (i.e., no additional benefit against hospitalization is assumed); this scenario causes the EJP to fall to $235.62 (23% reduction). A scenario performed from the societal cost perspective causes the EJP to increase slightly to $307.86 (1% increase). The productivity costs included in the societal perspective were $192,228,913 for the Stand-alone strategy and $176,722,406 for the Combination strategy, a savings of $15,506,507.

In two scenarios, the comparator was changed by substituting the base stand-alone vaccines used for alternatives. In a scenario where IIV-HD is used in place of IIV-Adj for influenza vaccination, the EJP increases to $353.66 (16% increase), while the use of BNT162b2 in place of mRNA-1273 causes the EJP to increase to $331.09 (9% increase).

Figure 4 shows a tornado diagram illustrating the impact of the remaining sensitivity analyses on the EJP. The additional protection that mRNA-1083 offers against hospitalization compared to mRNA-1273 impacts the EJP the most, followed by in-hospital mortality rates, waning of hospitalization VE, COVID-19 incidence, and percentage hospitalized with COVID-19. All other sensitivity analyses performed change the EJP by 3% or less.

**Figure 4.**
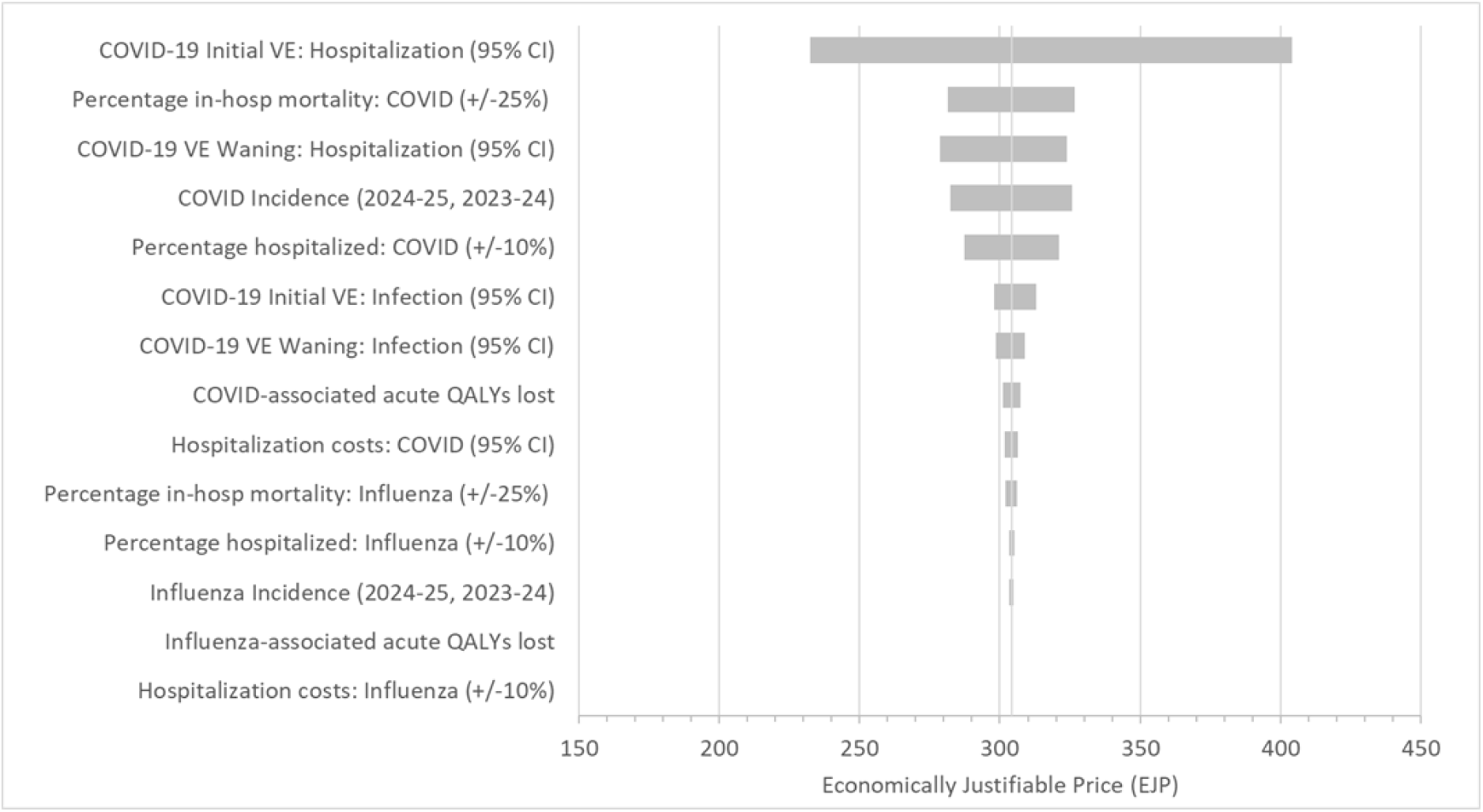
Sensitivity analysis: Impact of variation in model inputs on the estimated economically justifiable price for mRNA-1083 CI: confidence interval; In-hosp: in hospital; QALY: quality-adjusted life-years; VE: Vaccine effectiveness

## Discussion

This study estimated the population health and economic impacts of administering COVID-19 and influenza vaccinations separately (Stand-alone strategy) among adults aged ≥65 years in Canada, compared with integrating mRNA-1083 alongside these options (Combination strategy). The latter was projected to yield reductions in COVID-19 symptomatic infections (6%), hospitalizations (12%), and deaths (12%), and the corresponding influenza outcomes by approximately 2%. The reductions were driven by the increased vaccine coverage against COVID-19 and influenza that mRNA-1083 was assumed to generate and the higher VE of mRNA-1083 compared to mRNA-1273. The Combination strategy generated $222,470,201 in savings from COVID-19 and influenza-related treatment costs and $40,037,128 in savings from vaccine administration. The estimated EJP for mRNA-1083 is $304.11 at the $50,000 per QALY WTP. The value of mRNA-1083 is largely anchored to its capacity to increase vaccine coverage, with additional contributions from improved COVID-19 VE. Together, these factors reduced clinical and economic burden within the Combination strategy by enabling broader population protection, preventing infections, and improving vaccine delivery efficiency through fewer vaccine administrations.

Several scenario analyses were performed examining the impact of vaccine coverage and VE on the EJP, yielding a range of EJPs from $137.15 (if no rVE against COVID-19 for mRNA-1083 is assumed) to $328.48 (increased COVID-19 coverage and increased VE against influenza). Hospitalizations averted by the Combination strategy ranged from 0 to 6,237 for COVID-19 and 0 to 2,334 for influenza. Given the uncertainty associated with location of vaccine administration (i.e., physician office or pharmacy), several scenarios were performed to assess the impact of administration fees on the EJP, yielding a range from $283.80 (when the mRNA-1083 administration fee is increased to 2.5 times the base-case value) to $308.65 (when all vaccines are assumed to be administered in a physician’s office). The administration costs saved for each mRNA-1083 vaccine administered ranged from $7.74 to $14.22 per dose of mRNA-1083 administered, while the scenario assuming an increased administration fee for mRNA-1083 increased administration costs to $10.63 per dose. When the societal perspective was considered, there were $15,506,507 productivity loss costs saved with the Combination strategy. Model results are also sensitive to mRNA-1083 VE, in-hospital mortality rates, and COVID-19 incidence. In a scenario where IIV-HD is used in place of IIV-Adj for influenza vaccination, the EJP increases to $353.66 (16% increase), while the use of BNT162b2 in place of mRNA-1273 causes the EJP to increase to $331.09 (9% increase).

To our knowledge, this is the first study evaluating the value of a respiratory Combination vaccine in adults. The model structure aligns with published Canadian decision-analytic modelling studies, including the influenza model developed by the Quebec Immunization Committee (CIQ) [54,55] and the COVID-19 models by Miranda et al. [53] and Fust et al. [48]. Target populations between the CIQ model [54,55] and present study are similar, examining vaccination in individuals ≥65 years; further, key elements of the model, such as the inclusion of clinical outcomes such as hospitalizations, deaths, outpatient and emergency care are reflected in both studies to reflect the influenza public health burden. A full comparison with Miranda et al. [53] is provided in Fust et al. (2025) [48]; in brief, many COVID-related inputs were estimated for consistency with Miranda et al., and differences in results can be explained largely by differences in estimation of VE and the inclusion of medically attended cases only in the Miranda et al. publication (whereas the present study also includes those receiving no formal care). The present study was also designed so that the model would predict severe outcomes in alignment with hospitalization surveillance data for both influenza [3,61] and COVID [53,62], thus replicating observed hospitalization rates for both influenza and COVID-19 in Canada.

Jit et al. (2026) [28] provide a framework for evaluating the value of combination vaccines; based on this framework, the present study models operational efficiencies to the health system and more streamlined vaccine schedules, representing an important, although incomplete, estimation of combination vaccine value. Additionally, previously published literature has largely theorized the value of adult combination vaccines, described methodological frameworks[28], or estimated the value of pediatric combination vaccines. The present study provides a Canadian, adult-focused (≥65 years), public health impact and economic application of a combination vaccination strategy, and quantifies clinical outcomes, cost offsets, administration implications, and the resulting EJP.

Many of the limitations previously described in Fust et al. related to COVID-19 [48] also apply to this analysis. The mRNA-1083 VE is still being evaluated in clinical trials. For the purposes of this analysis, the mRNA-1083 VE for COVID-19 was assumed to be equivalent to mRNA-1283 and estimates for mRNA-1283 compared to mRNA-1273 and BNT162b2 need to be confirmed by head-to-head comparative effectiveness studies in the real-world setting. It was conservatively assumed that there would be no mRNA-1083 VE benefit against influenza infection or hospitalization, which likely underestimates the full benefits of mRNA-1083 vaccination. In clinical trials, mRNA-1010, the influenza component of mRNA-1083, has demonstrated non-inferiority to enhanced vaccines[97], as well as superiority to standard-dose vaccines (clinical trial P-304) [98]. mRNA-1083 also had a higher immune response against clinically relevant strains compared to high-dose influenza vaccines based on immunogenicity data in the P-301 clinical trial [34]. Additionally, no long-term costs of influenza or influenza-related complications included in the model, which may further underestimate the value of mRNA-1083.

The impact of mRNA-1083 on variables such as vaccine coverage and administration fees is highly uncertain. In this analysis, it was assumed that all individuals currently receiving both stand-alone vaccines at the same visit would switch to mRNA-1083, as these individuals have demonstrated acceptance of both antigens and comfort with coadministration. For these individuals, switching to a combination product likely does not change their behavioral burden, reduces injections, reduces administrative complexity and preserves the same clinical intent. The uptake for individuals currently receiving only one stand-alone vaccine (i.e., either COVID-19 or influenza) is less clear. It is also unclear how administration fees for combination vaccines such as mRNA-1083 may be implemented in clinical practice, and whether savings will be introduced via the reduction in injections, or if a premium administrative fee may be charged.

mRNA-1083 may reduce the clinical burden of COVID-19 and influenza in the NACI-recommended population (all aged ≥65), offering added benefit over current stand-alone COVID-19 and influenza vaccines in Canada. These benefits are driven by improvements in VE and increases in vaccine coverage through the streamlining of inefficiencies in existing routine vaccination programs. Health system savings with a combination vaccination strategy are also likely; however, these savings are dependent on the setting where combination vaccines are administered (physician’s office or pharmacy), the combination vaccine administrative fee and the unit cost of the new mRNA-1083 vaccine. However, results of this analysis suggest that a combination vaccination strategy provides good economic value when priced below $304.11 considering the $50,000 per QALY WTP threshold.

## Author Contributions

Model design: KF, MK, DM

Model programming: SC, KF

Model estimation: KF, MK, MB, DM

Analyses: KF, SC, MK, DM, MB

Writing: KF, MK, MB, DM

Critical review: MB, DM, NV

## Supporting information

Technical Appendix

## Data Availability

All data produced in the present work are contained in the manuscript.

## Acknowledgments

None

## Previous presentations

None

