## Supplementary material for "The Potential Clinical and Economic Impact of a Combination COVID-19 and Influenza Vaccine (mRNA-1083) in Canada": Technical Appendix

#### 1 Incidence of Influenza Infection

The approach for estimating incidence has been previously described in detail in Fust et al. (2025) [1]. The inputs used for the various steps in the estimation process are outlined here.

**Step 1**: Develop age-specific targets for the monthly hospitalization rates, by age group, for a one-year period

**Table 1. Target hospitalization rates per 100,000 for the calculation of infection incidence**

| Season | Sept | Oct | Nov | Dec | Jan | Feb | Mar | Apr | May | June | July | Aug |
|---|---|---|---|---|---|---|---|---|---|---|---|---|
| COVID-19 | | | | | | | | | | | | |
| 2023-2024[2] | 63.4 | 77.1 | 81.1 | 96.3 | 52.7 | 30.9 | 23.8 | 15.2 | 18.8 | 9.6 | 18.8 | 18.8 |
| 2024-2025[3] | 110.4 | 104.0 | 70.0 | 57.4 | 45.4 | 24.1 | 17.7 | 14.3 | 13.1 | 8.7 | 8.5 | 14.5 |
| Influenza | | | | | | | | | | | | |
| 2023-2024[4] | 1.5 | 3.8 | 32.0 | 91.6 | 38.8 | 10.5 | 8.5 | 5.9 | 3.8 | 1.5 | 1.0 | 0.1 |
| 2024-2025[5] | 1.0 | 2.1 | 3.5 | 29.3 | 42.0 | 80.6 | 103.6 | 43.8 | 8.9 | 1.6 | 1.2 | 0.1 |

**Step 2**: Enter age-specific probabilities of hospitalization (3.9% [COVID-19]; 9.09% [influenza]) [6-8]; in the absence of Canadian data, data from the United States Centers for Disease Control and Prevention (CDC) for influenza were used

**Step 3**: Enter assumed vaccination coverage (reflecting the 2023-2024 and 2024-2025 COVID-19 and influenza seasons)

Estimates of coverage for COVID-19 vaccines for the 2023-2024 and 2024-2025 seasons were obtained from Government of Canada data [9,10].

**Table 2. COVID-19 Base-case Coverage**

| | 2023-2024 | 2024-2025 |
|---|---|---|
| September | 7.9% | 8.6% |
| October | 23.3% | 25.2% |
| November | 38.1% | 41.3% |
| December | 41.0% | 44.5% |
| January | 42.6% | 46.2% |
| February | 44.0% | 47.7% |
| March | 45.4% | 49.2% |
| April | 46.7% | 50.6% |
| May | 48.0% | 52.1% |
| June | 49.4% | 53.5% |

| | 2023-2024 | 2024-2025 |
|---|---|---|
| July | 49.4% | 53.5% |
| August | 49.4% | 53.5% |

The Government of Canada Seasonal Influenza Vaccination Coverage reports [9,11] provide estimates of annual coverage for the 2023-2024 (72.7%) and 2024-2025 (63.2%) seasons for all adults ≥65 years. The model requires estimates of monthly coverage; the distribution of influenza vaccines, across months, was available from Public Health Agency of Canada data [12] (replicated in the table below); vaccination began in September 2023 and ended in January 2024, with the majority of vaccines provided in October and November. The coverage in each month was calculated by multiplying the annual coverage rate by the proportion in each month; cumulative coverage was subsequently calculated by summing the cumulative coverage in the previous month with the monthly coverage in the current month (Table 3). In the absence of data for 2024-2025 representing the monthly distribution of influenza vaccines, data from the 2023-2024 season was assumed to also apply to 2024-2025.

**Table 3. Calculation of Monthly Cumulative Coverage (Influenza)**

| | Proportion vaccinated in each month (%) | Monthly Coverage (%)* | | Cumulative Coverage (%)† | |
|---|---|---|---|---|---|
| | | 2023-2024 | 2024-2025 | 2023-2024 | 2024-2025 |
| September | 7.4% | 5.4% | 4.7% | 5.4% | 4.7% |
| October | 39.6% | 28.8% | 25.0% | 34.2% | 29.7% |
| November | 36.9% | 26.8% | 23.3% | 61.0% | 53.0% |
| December | 13.2% | 9.6% | 8.3% | 70.6% | 61.3% |
| January‡ | 3.0% | 2.1% | 1.9% | 72.7% | 63.2% |

*Calculated by multiplying annual coverage (72.7% for 2023-2024; 63.2% for 2024-2025) by the proportion vaccinated in each month [11,12]

†Calculated by summing the cumulative coverage in the previous month with the monthly coverage in the current month

‡All vaccinations were administered by January; accordingly, cumulative coverage from February– August is assumed equivalent to the January estimates

**Step 4**: Enter the assumed initial vaccine effectiveness (VE) against infection and hospitalization and the monthly linear waning rate over time.

*Influenza:* The 2023-2024 influenza VEs used during the calibration period were based on data published by Skowronski et al. (2024)[13] and represent the market mix of vaccines administered during the 2023-2024 influenza season. Skowronski et al. used a test-negative case-control design to estimate VE in those ages 20-64 years and ≥65 years receiving influenza vaccines beginning in October 2023 through January 2024 relative to individuals that did not receive the vaccine during the same time period. The primary VE outcome was medically attended outpatient acute respiratory illness due to laboratory-confirmed influenza. The adjusted (for age group, province, and calendar time) VE estimate against influenza A (the predominant strain during the study period, representing 94% of influenza positive specimens) was reported to be 70% (95% CI: 48%, 83%) for ages ≥65 years. Given that the VE estimates represent 3-month average VE values, the initial VE value for month 1 was back-calculated using a waning rate of 8%.

**Table 4. Vaccine Effectiveness Estimates Used in Incidence Calibration**

| Vaccine | Age Group | Infection | | Hospitalization | |
|---|---|---|---|---|---|
| | | Initial VE | Waning | Initial VE | Waning |
| Influenza | | | | | |
| 2023-2024 | 65+ years | 78% | 8% | 78% | 8% |
| 2024-2025 | 65+ years | 67% | 8% | 67% | 8% |
| COVID-19 | | | | | |
| 2023-2024 | 65+ | 40% | 4.75% | 63% | 2.5% |
| 2024-2025 | 65+ | 52% | 4.75% | 54% | 2.5% |

VE: Vaccine effectiveness

**Step 5:** Estimate the rates using model decision tree calculations

Base-case incidence values are displayed in Figure 1.

**Figure 1. COVID-19 incidence estimates**

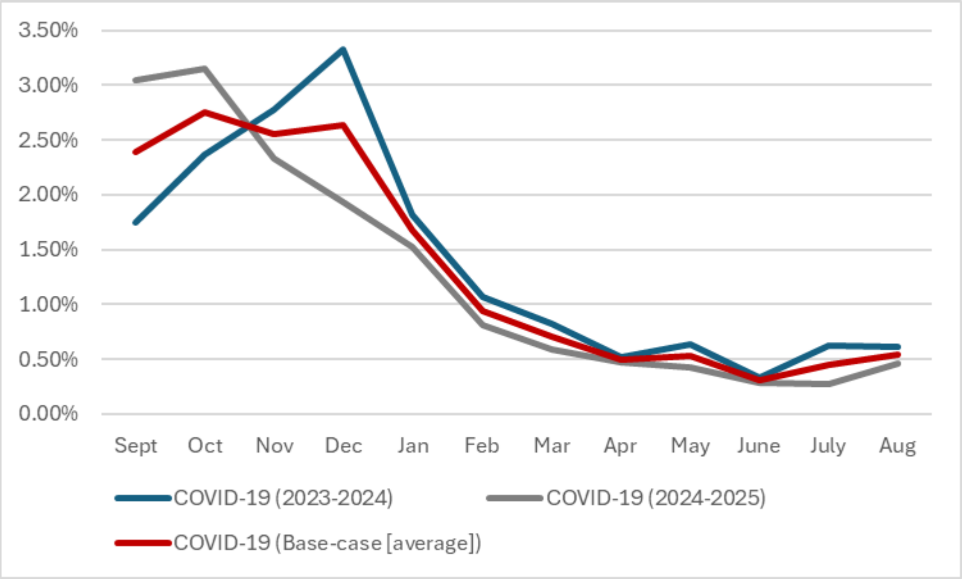

**Figure 2. Influenza incidence estimates**

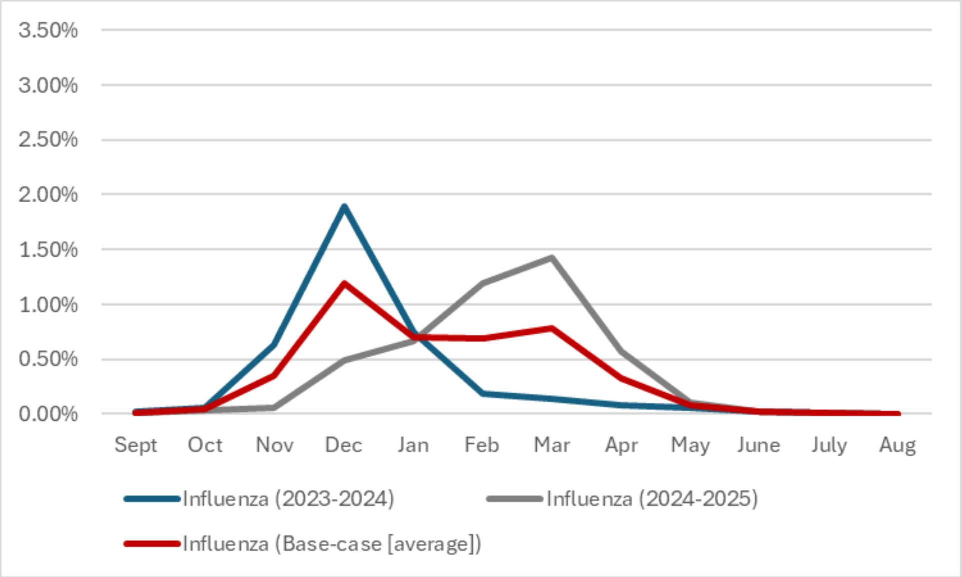

#### 2 Estimation of Base-Case Vaccine Coverage

The Government of Canada Seasonal Influenza Vaccination Coverage report [9] provides an estimate of annual coverage for all adults ≥65 years (63.2%) for the 2024-2025 season. The model requires estimates of monthly coverage; the distribution of influenza vaccines, across months, was available from Public Health Agency of Canada data [12] (replicated in the table below); vaccination began in September 2023 and ended in January 2024, with the majority of vaccines provided in October and November. It was assumed that this pattern would also apply to the 2024-2025 season. The coverage in each month was calculated by multiplying the annual coverage rate by the proportion in each month; cumulative coverage was subsequently calculated by summing the cumulative coverage in the previous month with the monthly coverage in the current month (Table 5).

**Table 5. Calculation of Base-Case Monthly Cumulative Coverage (2024-2025 Season)**

| | Proportion vaccinated in each month (%) | Monthly Coverage (%)* | Cumulative Coverage (%)† |
|---|---|---|---|
| September 2023 | 7.4% | 4.7% | 4.7% |
| October 2023 | 39.6% | 25.0% | 29.7% |
| November 2023 | 36.9% | 23.3% | 53.0% |
| December 2023 | 13.1% | 8.3% | 61.3% |
| January 2024‡ | 2.9% | 1.8% | 63.1% |

*Calculated by multiplying annual coverage (63.2% for ages 65+ years) by the proportion vaccinated in each month [9,12]

†Calculated by summing the cumulative coverage in the previous month with the monthly coverage in the current month

‡All vaccinations were administered by January; accordingly, cumulative coverage for February – August are assumed equivalent to the January estimates

Estimates of coverage for COVID-19 vaccines for the 2024-2025 season were calculated from Government of Canada data [9,10].

**Table 6. COVID-19 Base-case Coverage**

| | **2024-2025** |
|---|---|
| September | 8.6% |
| October | 25.2% |
| November | 41.3% |
| December | 44.5% |
| January | 46.2% |
| February | 47.7% |
| March | 49.2% |
| April | 50.6% |
| May | 52.1% |
| June | 53.5% |
| July | 53.5% |
| August | 53.5% |

#### 3 BNT162b2 Vaccine Efficacy

In a scenario analysis, BNT162b2 was used as the stand-alone COVID-19 vaccine in place of mRNA-1273.

As previously described in Fust et al. (2025), the vaccine efficacy for BNT-162b2 was calculated using the relative vaccine efficacy of mRNA-1283 compared to BNT-162b2 from an indirect treatment comparison [14,15]. Values are presented in the table below.

**Table 7. Initial vaccine effectiveness values (%) for scenario analysis (BNT162b2)**

| | mRNA-1283* | rVE of mRNA-1283 compared to BNT-162 | BNT-162b2 |
|---|---|---|---|
| **Symptomatic Infection** | 62.2% | 22.8 | 51.0% |
| **Hospitalization** | 73.3% | 44.1 | 52.1% |

*VE for mRNA-1083 assumed equal to mRNA-1283

#### 4 Cohort Size

**Table 8. Eligible Cohort Size - influenza**

| Age Group (Years) | Population Size | Percent Eligible* | Eligible Population Size | Source |
|---|---|---|---|---|
| 65+ | 8,365,300 | 100% | 8,365,300 | Statistics Canada [16] |

### 5 Adverse Event Rates

#### Table 9. Adverse Event Rates

| | | Standalone COVID-19 Vaccines | | Standalone Influenza Vaccines | | |
|---|---|---|---|---|---|---|
| | **mRNA-1083*** | **mRNA-1273** | **BNT162b2**** | **IIV3-Adj†** | **TIV-HD** | **Source** |
| Grade 3 Local | 2.2% | 0.1% | 0.1% | 1.0% | 1.0% | Rudman Spergel et al. (2025) [17] |
| Grade 3 Systemic | 7.6% | 1.6% | 1.6% | 1.7% | 1.7% | Rudman Spergel et al. (2025) [17] |
| Grade 4 Local | 0.0% | 0.0% | 0.0% | 0.0% | 0.0% | Rudman Spergel et al. (2025) [17] |
| Grade 4 Systemic‡ | 0.1% | 0.1% | 0.0% | 0.1% | 0.1% | Rudman Spergel et al. (2025) [17] |
| Anaphylaxis | 0.0005% | 0.0005% | 0.0005% | 0.0005% | 0.0005% | Klein et al. (2021)[18] |
| Guillain-Barré Syndrome | 0.0% | 0.0% | 0.0% | 0.00007% | 0.00007% | Perez-Vilar (2021)[19] |

*Estimates for mRNA-1083 were obtained from the mRNA-1083 clinical trial (Rudman Spergel et al. 2025); estimates may differ from published rates because they were obtained from a separate analysis based on the clinical trial data (Table 14.3.1.2.5a from the Clinical Study Report) that provides estimates for the comparators (TIV-HD and mRNA-1273) separately

**Assumed equal to mRNA-1273 (with no grade 4 events)

†Assumed equal to TIV-HD

‡Grade 4 Systemic AE rates were calculated by assuming the same ratio as observed between TIV-HD & mRNA-1273 for Grade 3 Local AEs

#### 6 Lost Productivity

Lost productivity-related inputs are included in analyses performed from the societal cost perspective only and are presented in the table below.

**Table 10. Lost Productivity Inputs**

| Model Parameter | Value | Data Source |
|---|---|---|
| Labour force participation rate[a] | | |
| 65-79 | 17.3% | Statistics Canada [20] |
| 80-100 | 8.4% | Statistics Canada [20] |
| Daily wage[a] | $265.95 | Statistics Canada [21] |
| Time loss (days) due to: | | |
| Vaccination (COVID-19 or influenza) | 0.06 | Miranda 2024 [22] |
| Adverse events | | |
| Grade 3 Local | 0.19 | Rousculp 2024 [23] |
| Grade 3 Systemic | 0.19 | Rousculp 2024 [23] |
| Anaphylaxis | 5.86 | CIHI [24] |
| Myocardisit/pericarditis | 3.67 | CIHI [24] |
| GBS | 6.83 | CMDB [25]<br>CIHI 2022 [26]<br>PLPB [27] |
| Infection-related myocarditis | 3.67 | CIHI [24] |
| COVID-19 infection-related: | | |
| No formal care | 0 | Assumption |
| Outpatient care | 3.57 | Miranda 2024 [22] |
| Hospitalization: | | |
| No ICU or Ventilator | 18.57 | CIHI [24] |
| ICU Only | 25.29 | CIHI [24] |
| ICU with Ventilator | 25.29 | CIHI [24] |
| Post-acute care (outpatient/hospitalized) | 23.3 | Miranda 2024 [22] |
| Influenza infection-related: | | |
| No formal care | 0 | Assumption |
| Outpatient care only | 5.23 | Derived from Waite 2022 [28] |
| Hospitalization* | 5.93 | Derived from Waite 2022 [28] |

CMDB, Canadian MIS Database; CIHI, Canadian Institute for Health Information; PLPB, Patient Level Physician Billing repository.

[a]updated from Fust et al. (2025) [1] for COVID-19 component

*Hospitalized patients are also applied time loss associated with outpatient care, to reflect days with symptoms prior to hospitalization occurring

#### 7 Additional Base-Case Results

**Table 11. Base-Case Results: QALYs Lost**

| | **Stand-Alone Strategy** | | **Combination Strategy** | | **Difference (Combination - Stand-Alone Strategy)** | | **Percent Change** | |
|---|---|---|---|---|---|---|---|---|
| | **COVID-19** | **Influenza** | **COVID-19** | **Influenza** | **COVID-19** | **Influenza** | **COVID-19** | **Influenza** |
| **QALYs Lost (Death)** | **78,897** | **44,677** | **69,492** | **43,984** | **-9,405** | **-693** | **-12%** | **-2%** |
| **QALYs Lost (Morbidity)** | | | | | | | | |
| No formal care cases (including post-infection) | 3,033 | 2,602 | 2,842 | 2,562 | -192 | -40 | -6% | -2% |
| Outpatient care cases (including post-infection) | 11,856 | 4,175 | 11,172 | 4,110 | -683 | -65 | -6% | -2% |
| Hospitalizations (including post-discharge) | 4,358 | 678 | 3,839 | 667 | -520 | -11 | -12% | -2% |
| COVID 19 infection-related myocarditis cases | 4 | -- | 4 | -- | 0 | -- | -6% | -- |
| ***Sub-total*** | **98,148** | **52,132** | **87,349** | **51,324** | **-10,799** | **-808** | **-11%** | **-2%** |
| **QALYs Lost (Vaccine-related Adverse Events)** | **127** | | **227** | | **100** | | **79%** | |

**Table 12. Base-Case Results: Adverse Events**

| Event | Stand-Alone Strategy | | Combination Strategy | | | Difference (Combination - Stand-Alone Strategy) | Percent Change |
|---|---|---|---|---|---|---|---|
| | mRNA-1273 | IIV3-Adj | mRNA-1083 | mRNA-1273 | IIV3-Adj | | |
| Grade 3 Local | 42,517 | 52,869 | 90,478 | 11,179 | 14,026 | 20,297 | 21% |
| Grade 4 Local | 0 | 0 | 0 | 0 | 0 | 0 | - |
| Grade 3 Systemic | 73,041 | 90,826 | 312,561 | 19,204 | 24,095 | 191,993 | 117% |
| Grade 4 Systemic | 2,180 | 2,711 | 6,169 | 573 | 719 | 2,570 | 53% |
| Myocarditis | 0 | -- | 0 | 0 | -- | 0 | - |
| Anaphylaxis | 22 | 26 | 20 | 6 | 7 | -15 | -31% |
| Guillan-Barré Syndrome | 0 | 4 | 0 | 0 | 1 | -3 | -73% |

**Table 13. Scenario and Sensitivity Analysis Results: Change (Δ) in Clinical Outcomes (Combination Strategy – Stand-alone Strategy)**

| | **COVID-19** | | | | | | **Influenza** | | | | | |
|---|---|---|---|---|---|---|---|---|---|---|---|---|
| **Scenario** | **Δ in Inf.** | **% Δ from Base-Case** | **Δ in Hosp.** | **% Δ from Base-Case** | **Δ in Deaths** | **% Δ from Base-Case** | **Δ in Inf.** | **% Δ from Base-Case** | **Δ in Hosp.** | **% Δ from Base-Case** | **Δ in Deaths** | **% Δ from Base-Case** |
| **Base Case** | **-71,074** | **--** | **-5,008** | **--** | **-935** | **--** | **-3,985** | **--** | **-362** | **--** | **-69** | **--** |
| **Coverage Scenarios** | | | | | | | | | | | | |
| Increased COVID-19 VCR | -94,688 | 33.2% | -6,237 | 24.5% | -1,164 | 24.5% | -3,979 | -0.2% | -362 | -0.2% | -69 | -0.2% |
| No increase in COVID-19 or influenza VCR | -21,823 | -69.3% | -2,334 | -53.4% | -436 | -53.4% | 7 | -100.2% | 1 | -100.2% | 0 | -100.2% |
| No rVE against COVID-19 | -41,550 | -41.5% | -1,851 | -63.0% | -345 | -63.0% | -3,994 | 0.2% | -363 | 0.2% | -69 | 0.2% |
| No rVE against COVID-19, increased COVID-19 VCR | -62,327 | -12.3% | -2,776 | -44.6% | -518 | -44.6% | -3,988 | 0.1% | -363 | 0.1% | -69 | 0.1% |
| No rVE against COVID-19, no increase in COVID-19 or influenza VCR | 0 | -100% | 0 | -100% | 0 | -100.0% | 0 | -100% | 0 | -100% | 0 | -100% |
| Include increased rVEs against influenza (due to egg adaptation) | -71,072 | 0.0% | -5,008 | 0.0% | -935 | 0.0% | -7,449 | 86.9% | -677 | 86.9% | -129 | 86.9% |
| Include increased rVEs against influenza (due to egg adaptation), increased COVID-19 VCR | -94,686 | 33.2% | -6,237 | 24.5% | -1,164 | 24.5% | -7,792 | 95.5% | -708 | 95.5% | -135 | 95.5% |
| Include increased rVEs against influenza (due to egg adaptation), no increase in COVID-19 or influenza VCR | -21,821 | -69.3% | -2,334 | -53.4% | -436 | -53.4% | -2,545 | -36.1% | -231 | -36.1% | -44 | -36.1% |
| **Vaccine Effectiveness Scenarios** | | | | | | | | | | | | |
| COVID-19 rVE hosp. = rVE inf. | -71,086 | 0.02% | -2,974 | -40.6% | -555 | -40.6% | -3,991 | 0.1% | -363 | 0.1% | -69 | 0.1% |
| **Comparator Scenarios** | | | | | | | | | | | | |

| | COVID-19 | | | | | | Influenza | | | | | |
|---|---|---|---|---|---|---|---|---|---|---|---|---|
| **Scenario** | **Δ in Inf.** | **% Δ from Base-Case** | **Δ in Hosp.** | **% Δ from Base-Case** | **Δ in Deaths** | **% Δ from Base-Case** | **Δ in Inf.** | **% Δ from Base-Case** | **Δ in Hosp.** | **% Δ from Base-Case** | **Δ in Deaths** | **% Δ from Base-Case** |
| TIV-HD replaces IIV3-Adj | -71,074 | 0.0% | -5,008 | 0.0% | -935 | 0.0% | -3,985 | 0.0% | -362 | 0.0% | -69 | 0.0% |
| BNT-162b2 replaces mRNA-1273 | -92,182 | 29.7% | -5,736 | 14.5% | -1,071 | 14.5% | -3,983 | -0.1% | -362 | -0.1% | -69 | -0.1% |
| **Incidence Scenarios** | | | | | | | | | | | | |
| COVID-19 Incidence: 2023-2024 | -79,290 | 11.6% | -5,599 | 11.8% | -1,045 | 11.8% | -3,983 | -0.04% | -362 | -0.04% | -69 | -0.04% |
| COVID-19 Incidence: 2024-2025 | -62,858 | -11.6% | -4,417 | -11.8% | -824 | -11.8% | -3,987 | 0.04% | -362 | 0.04% | -69 | 0.04% |
| Influenza Incidence: 2023-24 | -71,072 | 0.0% | -5,008 | 0.0% | -935 | 0.0% | -4,319 | 8.4% | -393 | 8.4% | -75 | 8.4% |
| Influenza Incidence: 2024-25 | -71,076 | 0.0% | -5,008 | 0.0% | -935 | 0.0% | -3,652 | -8.4% | -332 | -8.4% | -63 | -8.4% |
| **Deterministic Sensitivity Analyses** | | | | | | | | | | | | |
| COVID-19 Initial VE: Infection lower bound | -41,529 | -41.6% | -5,008 | 0.0% | -935 | 0.0% | -3,985 | 0.0% | -362 | 0.0% | -69 | 0.0% |
| COVID-19 Initial VE: Infection upper bound | -113,865 | 60.2% | -5,008 | 0.0% | -935 | 0.0% | -3,985 | 0.0% | -362 | 0.0% | -69 | 0.0% |
| COVID-19 Initial VE: Hosp lower bound | -71,087 | 0.02% | -2,876 | -42.6% | -537 | -42.6% | -3,991 | 0.2% | -363 | 0.2% | -69 | 0.2% |
| COVID-19 Initial VE: Hosp upper bound | -71,056 | -0.03% | -7,977 | 59.3% | -1,489 | 59.3% | -3,976 | -0.2% | -361 | -0.2% | -69 | -0.2% |
| COVID-19 VE Waning: Infection lower bound | -94,681 | 33.2% | -5,008 | 0.0% | -935 | 0.0% | -3,985 | 0.0% | -362 | 0.0% | -69 | 0.0% |
| COVID-19 VE Waning: Infection upper bound | -44,183 | -37.8% | -5,008 | 0.0% | -935 | 0.0% | -3,985 | 0.0% | -362 | 0.0% | -69 | 0.0% |
| COVID-19 VE Waning: Hosp lower bound | -71,072 | 0.00% | -5,593 | 11.69% | -1,044 | 11.69% | -3,984 | -0.02% | -362 | -0.02% | -69 | -0.02% |
| COVID-19 VE Waning: Hosp upper bound | -71,077 | 0.0% | -4,256 | -15.0% | -794 | -15.0% | -3,986 | 0.02% | -362 | 0.02% | -69 | 0.02% |
| Percentage hospitalized (COVID) -10% | -71,079 | 0.01% | -4,507 | -10.0% | -841 | -10.0% | -3,987 | 0.05% | -362 | 0.05% | -69 | 0.05% |

| | COVID-19 | | | | | | Influenza | | | | | |
|---|---|---|---|---|---|---|---|---|---|---|---|---|
| **Scenario** | **Δ in Inf.** | **% Δ from Base-Case** | **Δ in Hosp.** | **% Δ from Base-Case** | **Δ in Deaths** | **% Δ from Base-Case** | **Δ in Inf.** | **% Δ from Base-Case** | **Δ in Hosp.** | **% Δ from Base-Case** | **Δ in Deaths** | **% Δ from Base-Case** |
| Percentage hospitalized (COVID) +10% | -71,069 | -0.01% | -5,509 | 10.0% | -1,028 | 10.0% | -3,983 | -0.05% | -362 | -0.05% | -69 | -0.05% |
| Percentage hosp. (Influenza) -10% | -71,075 | 0.0% | -5,008 | 0.0% | -935 | 0.0% | -3,985 | 0.0% | -326 | -10.0% | -62 | -10.0% |
| Percentage hosp. (Influenza) +10% | -71,073 | 0.0% | -5,008 | 0.0% | -935 | 0.0% | -3,985 | 0.0% | -398 | 10.0% | -76 | 10.0% |
| Percentage in-hosp mortality (COVID) -25% | -71,084 | 0.01% | -5,008 | 0.01% | -747 | -20.1% | -3,990 | 0.1% | -363 | 0.1% | -69 | 0.1% |
| Percentage in-hosp mortality (COVID) +25% | -71,064 | -0.01% | -5,008 | -0.01% | -1,122 | 20.1% | -3,981 | -0.1% | -362 | -0.1% | -69 | -0.1% |
| Percentage in-hosp mortality (Influenza) -25% | -71,077 | 0.0% | -5,008 | 0.0% | -935 | 0.0% | -3,986 | 0.01% | -362 | 0.01% | -52 | -25.0% |
| Percentage in-hosp mortality (Influenza) +25% | -71,071 | 0.0% | -5,008 | 0.0% | -935 | 0.0% | -3,985 | -0.01% | -362 | -0.01% | -86 | 25.0% |

Δ: Change; Inf.: Infection; Hosp.: Hospitalization; VE: Vaccine effectiveness

**Table 14. Scenario and Sensitivity Analysis Results: Economically Justifiable Price**

| **Scenario** | **Economically Justifiable Price** | **% Change from Base Case** |
|---|---|---|
| **Base case** | **$304.11** | **--** |
| **Societal Cost Perspective** | $307.86 | 1.2% |
| **Comparator Scenarios** | | |
| TIV-HD replaces IIV3-Adj | $353.66 | 16.3% |
| BNT-162b2 replaces mRNA-1273 | $331.09 | 8.9% |
| **Vaccine Effectiveness Scenarios** | | |
| COVID-19 rVE hospitalization = rVE infection | $235.62 | -22.5% |
| **Incidence Scenarios** | | |
| COVID-19 Incidence: 2023-2024 | $325.71 | 7.1% |
| COVID-19 Incidence: 2024-2025 | $282.50 | -7.1% |
| Influenza Incidence: 2023-24 | $305.00 | 0.3% |
| Influenza Incidence: 2024-25 | $303.21 | -0.3% |
| **Deterministic Sensitivity Analyses** | | |
| COVID-19 Initial VE: Infection lower bound | $297.96 | -2.0% |
| COVID-19 Initial VE: Infection upper bound | $313.01 | 2.9% |
| COVID-19 Initial VE: Hosp lower bound | $232.34 | -23.6% |
| COVID-19 Initial VE: Hosp upper bound | $404.08 | 32.9% |
| COVID-19 VE Waning: Infection lower bound | $309.02 | 1.6% |
| COVID-19 VE Waning: Infection upper bound | $298.51 | -1.8% |
| COVID-19 VE Waning: Hosp lower bound | $323.82 | 6.5% |
| COVID-19 VE Waning: Hosp upper bound | $278.77 | -8.3% |
| Percentage hospitalized (COVID) -10% | $287.25 | -5.5% |
| Percentage hospitalized (COVID) +10% | $320.96 | 5.5% |
| Percentage hospitalized (Influenza) -10% | $303.19 | -0.3% |
| Percentage hospitalized (Influenza) +10% | $305.02 | 0.3% |
| Percentage in-hospital mortality (COVID) -25% | $281.53 | -7.4% |
| Percentage in- hospital mortality (COVID) +25% | $326.68 | 7.4% |
| Percentage in- hospital mortality (Influenza) -25% | $302.02 | -0.7% |
| Percentage in- hospital mortality (Influenza) +25% | $306.19 | 0.7% |
| Hospitalization costs (COVID) lower 95%CI | $301.71 | -0.8% |
| Hospitalization costs (COVID) upper 95%CI | $306.50 | 0.8% |
| Hospitalization costs (Influenza) -10% | $304.02 | 0.0% |
| Hospitalization costs (Influenza) +10% | $304.19 | 0.0% |
| COVID-associated acute QALYs lost (lower bound) | $301.02 | -1.0% |
| COVID-associated acute QALYs lost (upper bound) | $307.21 | 1.0% |
| Influenza -associated acute QALYs lost (lower bound) | $303.82 | -0.1% |
| Influenza -associated acute QALYs lost (upper bound) | $304.40 | 0.1% |

**Table 15. Number of Doses (Base-Case and Coverage Scenarios)**

| | Coverage Scenarios | | |
|---|---|---|---|
| | **Base-Case (10% Increase COVID-19 VCR; 3% Increase Influenza VCR)** | **Increased COVID-19 VCR (15% Increase COVID-19 VCR; 3% Increase Influenza VCR)** | **No Increase in VCR (0% Increase COVID-19 VCR; 0% Increase Influenza VCR)** |
| **Total Stand-Alone Strategy** | | | |
| mRNA-1273 | 4,475,436 | 4,475,436 | 4,475,436 |
| IIV3-Adj | 5,286,870 | 5,286,870 | 5,286,870 |
| **Total Combination Strategy** | | | |
| mRNA-1273 | 1,176,705 | 1,176,705 | 1,427,664 |
| IIV3-Adj | 1,402,568 | 984,303 | 2,239,098 |
| mRNA-1083 | 4,135,261 | 4,553,526 | 3,047,772 |

VCR: Vaccine coverage rate

#### 8 CHEERS Checklist

| | Item | Guidance for Reporting | Reported in section |
|---|---|---|---|
| **TITLE** | | | |
| Title | 1 | Identify the study as an economic evaluation and specify the interventions being compared. | Title Page |
| **ABSTRACT** | | | |
| Abstract | 2 | Provide a structured summary that highlights context, key methods, results and alternative analyses. | Abstract |
| **INTRODUCTION** | | | |
| Background and objectives | 3 | Give the context for the study, the study question and its practical relevance for decision making in policy or practice. | Introduction |
| **METHODS** | | | |
| Health economic analysis plan | 4 | Indicate whether a health economic analysis plan was developed and where available. | Analysis plan not developed |
| Study population | 5 | Describe characteristics of the study population (such as age range, demographics, socioeconomic, or clinical characteristics). | Introduction; Methods Overview |
| Setting and location | 6 | Provide relevant contextual information that may influence findings. | Overview |
| Comparators | 7 | Describe the interventions or strategies being compared and why chosen. | Overview |
| Perspective | 8 | State the perspective(s) adopted by the study and why chosen. | Overview |
| Time horizon | 9 | State the time horizon for the study and why appropriate. | Overview |
| Discount rate | 10 | Report the discount rate(s) and reason chosen. | Quality of Life |
| Selection of outcomes | 11 | Describe what outcomes were used as the measure(s) of benefit(s) and harm(s). | Overview; Model Structure |
| Measurement of outcomes | 12 | Describe how outcomes used to capture benefit(s) and harm(s) were measured. | Vaccine Coverage; Vaccine Effectiveness; Probabilities |
| Valuation of outcomes | 13 | Describe the population and methods used to measure and value outcomes. | Target Population |
| Measurement and valuation of resources | 14 | Describe how costs were valued. | Healthcare costs; Lost productivity |

| | Item | Guidance for Reporting | Reported in section |
|---|---|---|---|
| and costs | | | (Technical Appendix) |
| Currency, price date, and conversion | 15 | Report the dates of the estimated resource quantities and unit costs, plus the currency and year of conversion. | Healthcare costs |
| Rationale and description of model | 16 | If modelling is used, describe in detail and why used. Report if the model is publicly available and where it can be accessed. | Overview; Model Structure |
| Analytics and assumptions | 17 | Describe any methods for analysing or statistically transforming data, any extrapolation methods, and approaches for validating any model used. | Methods |
| Characterizing heterogeneity | 18 | Describe any methods used for estimating how the results of the study vary for sub-groups. | Not applicable |
| Characterizing distributional effects | 19 | Describe how impacts are distributed across different individuals or adjustments made to reflect priority populations. | Not applicable |
| Characterizing uncertainty | 20 | Describe methods to characterize any sources of uncertainty in the analysis. | Sensitivity Analyses; Scenario Analyses |
| Approach to engagement with patients and others affected by the study | 21 | Describe any approaches to engage patients or service recipients, the general public, communities, or stakeholders (e.g., clinicians or payers) in the design of the study. | Not applicable |
| **RESULTS** | | | |
| Study parameters | 22 | Report all analytic inputs (e.g., values, ranges, references) including uncertainty or distributional assumptions. | Included in Methods/Tech appendix & DSA/Scenario Results |
| Summary of main results | 23 | Report the mean values for the main categories of costs and outcomes of interest and summarise them in the most appropriate overall measure. | Results |
| Effect of uncertainty | 24 | Describe how uncertainty about analytic judgments, inputs, or projections affect findings. Report the effect of choice of discount rate and time horizon, if applicable. | Deterministic Sensitivity Analyses; Scenario Analyses |
| Effect of engagement with patients and others affected by the study | 25 | Report on any difference patient/service recipient, general public, community, or stakeholder involvement made to the approach or findings of the study | Not Applicable |

| | Item | Guidance for Reporting | Reported in section |
|---|---|---|---|
| **DISCUSSION** | | | |
| Study findings, limitations, generalizability, and current knowledge | 26 | Report key findings, limitations, ethical or equity considerations not captured, and how these could impact patients, policy, or practice. | Discussion |
| **OTHER RELEVANT INFORMATION** | | | |
| Source of funding | 27 | Describe how the study was funded and any role of the funder in the identification, design, conduct, and reporting of the analysis | Transparency |
| Conflicts of interest | 28 | Report authors conflicts of interest according to journal or International Committee of Medical Journal Editors requirements. | Transparency |

[10] Government of Canada. COVID-19 Canadian Respiratory Virus Surveillance Report 2025 [cited 2026 March].

[24] Canadian Institute for Health Information. Patient Cost Estimator Results. . Ottawa, Ontario, Canada2024.

[25] Canadian Institute for Health Information. Canadian MIS Database (CMDB), Discharge Abstract Database and Hospital Morbidity Database. 2017–2018 to 2021–2022.

[26] Canadian Institute for Health Information. CMG+ grouping methodology. 2022.

[27] Canadian Institute for Health Information. Patient Level Physician Billing repository (PLPB), Discharge Abstract Database and Hospital Morbidity Database,. 2017–2018 to 2021–2022.

[28] Waite NM, Pereira JA, Houle SKD, et al. The impact of influenza on the ability to work, volunteer and provide care: results from an online survey of Canadian adults 50 years and older. BMC Public Health. 2022;22(1):2119.
